# Machine learning predicts liver cancer risk from routine clinical data: a large population-based multicentric study

**DOI:** 10.1101/2024.11.03.24316662

**Authors:** Jan Clusmann, Paul-Henry Koop, David Y. Zhang, Felix van Haag, Omar S. M. El Nahhas, Tobias Seibel, Laura Žigutytė, Apichat Kaewdech, Julien Calderaro, Frank Tacke, Tom Luedde, Daniel Truhn, Tony Bruns, Kai Markus Schneider, Jakob N. Kather, Carolin V. Schneider

**Affiliations:** Department of Medicine III, University Hospital RWTH Aachen, Aachen, Germany; Else Kroener Fresenius Center for Digital Health, Technical University Dresden, Dresden, Germany; Department of Genetics, Perelman School of Medicine, University of Pennsylvania, Philadelphia, USA; Department of Medicine, Perelman School of Medicine, University of Pennsylvania, Philadelphia, USA; StratifAI GmbH, Dresden, Germany; Gastroenterology and Hepatology Unit, Division of Internal Medicine, Faculty of Medicine, Prince of Songkla University, Songkhla 90110, Thailand; Université Paris Est Créteil, INSERM, IMRB, F-94010, Créteil, France; Assistance Publique-Hôpitaux de Paris, Henri Mondor-Albert Chenevier University Hospital, Department of Pathology, Créteil, France; Inserm, U955, Team 18, Créteil, France; European Reference Network (ERN) RARE-LIVER, Créteil, France; Department of Hepatology and Gastroenterology, Charité - Universitätsmedizin Berlin, Campus Virchow-Klinikum and Campus Charité Mitte, Berlin, Germany; Department for Gastroenterology, Hepatology and Infectiology, University Hospital Düsseldorf, Germany; Department of Diagnostic and Interventional Radiology, University Hospital Aachen, Germany; Department of Medicine I, University Hospital Dresden, Dresden, Germany; Center for Regenerative Therapies Dresden (CRTD), Technische Universität (TU), Dresden, Germany; Department of Medical Oncology, National Center for Tumor Diseases (NCT), Heidelberg University Hospital, Heidel- berg, Germany

**Keywords:** Machine learning, hepatocellular carcinoma, risk stratification, early detection, big data, population-cohort, prediction, screening

## Abstract

**Background and aims:** Hepatocellular carcinoma (HCC) is a highly fatal tumor, for which early detection and risk stratification is crucial, yet remains challenging. We aimed to develop an interpretable machine-learning framework for HCC risk stratification based on routinely collected clinical data.

**Methods:** We leverage data obtained from over 900,000 individuals and 983 cases of HCC across two large-scale population-based cohorts: the UK Biobank study and the “All Of Us Research Program”. For all of these patients, clinical data from timepoints years before diagnosis of HCC was available. We integrate data modalities including demographics, electronic health records, lifestyle, routine blood tests, genomics and metabolomics to offer a unique, multi-modal perspective on HCC risk.

**Results:** Our random-forest-based model significantly outperforms all publicly available state-of-the-art risk-scores, with an AUROC of 0.88 both for internal and external test sets. We demonstrate robustness of our model across ethnic subgroups, a major advance over previous models with variable performance by ethnicity. Further, we perform extensive feature-importance analysis, showcasing our approach as an interpretable framework. We provide all model weights and an open-source web calculator to facilitate further validation of our model.

**Conclusion:** Our study presents a robust and interpretable machine-learning framework for HCC risk stratification, which offers the potential to improve early detection and could ultimately reduce disease burden through targeted interventions.

**Lay summary:** Finding liver cancer early is crucial for successful treatment. Therefore, screening with abdominal ultrasound can be performed. However, it is not clear who should receive ultrasound screening, as with the current standard of screening only patients with liver cirrhosis, a severe liver disease, many patients are diagnosed with liver cancer in late stages. Therefore, we trained a machine learning model, acting like many decision trees at the same time, to detect patients with high risk of liver cancer by looking at patterns of almost 1000 cases of liver cancer in a population of 900.000 individuals. In a separate set of patients, which the model has not seen during training, our model worked better than all available models. Additionally, we investigated 1. how the model comes to its prediction, 2. whether it works in males and females alike and 3. which data is most relevant for the model. Like this, our model can help sort patients into categories like “high-risk”, “medium-risk” and “low-risk”, via which screening strategies can then be decided, to help improve early detection of liver cancer.

## Introduction

Hepatocellular carcinoma (HCC) is the fifth most common malignancy and third leading cause of cancer-associated death. Its incidence has tremendously increased in the last years, highlighting HCC as a major public health concern, especially in structurally disadvantaged regions, locally and globally ^1–3^.

Current screening protocols for HCC predominantly target patients already diagnosed with liver cirrhosis and rely heavily on resource-intensive imaging technologies, which are not universally accessible. Despite screening, the majority of HCC cases are diagnosed only at late disease stages, drastically worsening prognosis ^4^. Additionally, these protocols fail to consider multifactorial risk factors such as lifestyle, pre-existing health conditions, blood tests or omics signatures, which could identify a broader at-risk population ^5–8^. This gap is particularly critical as the prevalence of metabolic dysfunction-associated steatotic liver disease (MASLD) and its related HCC cases continue to rise ^9^. This highlights the need for a screening strategy that is more affordable, more inclusive and more efficient at detecting patients with high risk of HCC. Imaging-based screening could then be specifically targeted at this target group.

Integrating the vast array of patient data to assess disease risk represents a significant challenge in modern medicine, particularly as screening is typically conducted by general practitioners who must possess a broad knowledge base across a wide range of diseases, each with its own set of unique risk factors. Machine learning (ML) algorithms present a promising solution to this challenge, especially for HCC, and could further reduce healthcare disparities, if developed cautiously ^10^. Several studies have shown the superiority of ML algorithms over traditional regression analyses or few-parameter based decision-support systems as implemented in current guidelines ^1,11–17^. However, a major criticism is the lack of generalizability due to small cohorts, lack of published model weights, a lack of external testing ^18^, as well as usage of highly curated, retrospective datasets that do not necessarily represent real-world scenarios.

In response to these challenges, we developed a novel, non-invasive multimodal screening tool that integrates these diverse risk factors into a comprehensive risk stratification model. Our study uses the extensive data available from the UK Biobank (UKB) and the “All Of Us Research program” (AOU), both of which include information extensive questionnaires, electronic health records, blood parameters and genomics for each over 500,000 participants, with ongoing follow-up since 2006 for UKB ^19–22^ and extensive retrospective documentation for AO<U ^23,24^. By reducing reliance on previously diagnosed cirrhosis and instead, including a wider array of risk determinants, this tool aims to improve early detection of HCC, particularly in patients who may benefit from regular screening but are currently overlooked. This approach not only promises to expand the efficacy of current screening practices but also extends its reach to under-resourced regions, potentially transforming HCC prognosis on a global scale.

Our study specifically investigates the individual and synergistic contributions of various data modalities, such as genotypic information versus lifestyle and blood data, to highlight the distinct advantages and augmented value each brings to enhancing HCC risk stratification. We hereby especially emphasize current underutilization of more accessible and cost-effective data types (Figure 1).

**Fig. 1:**
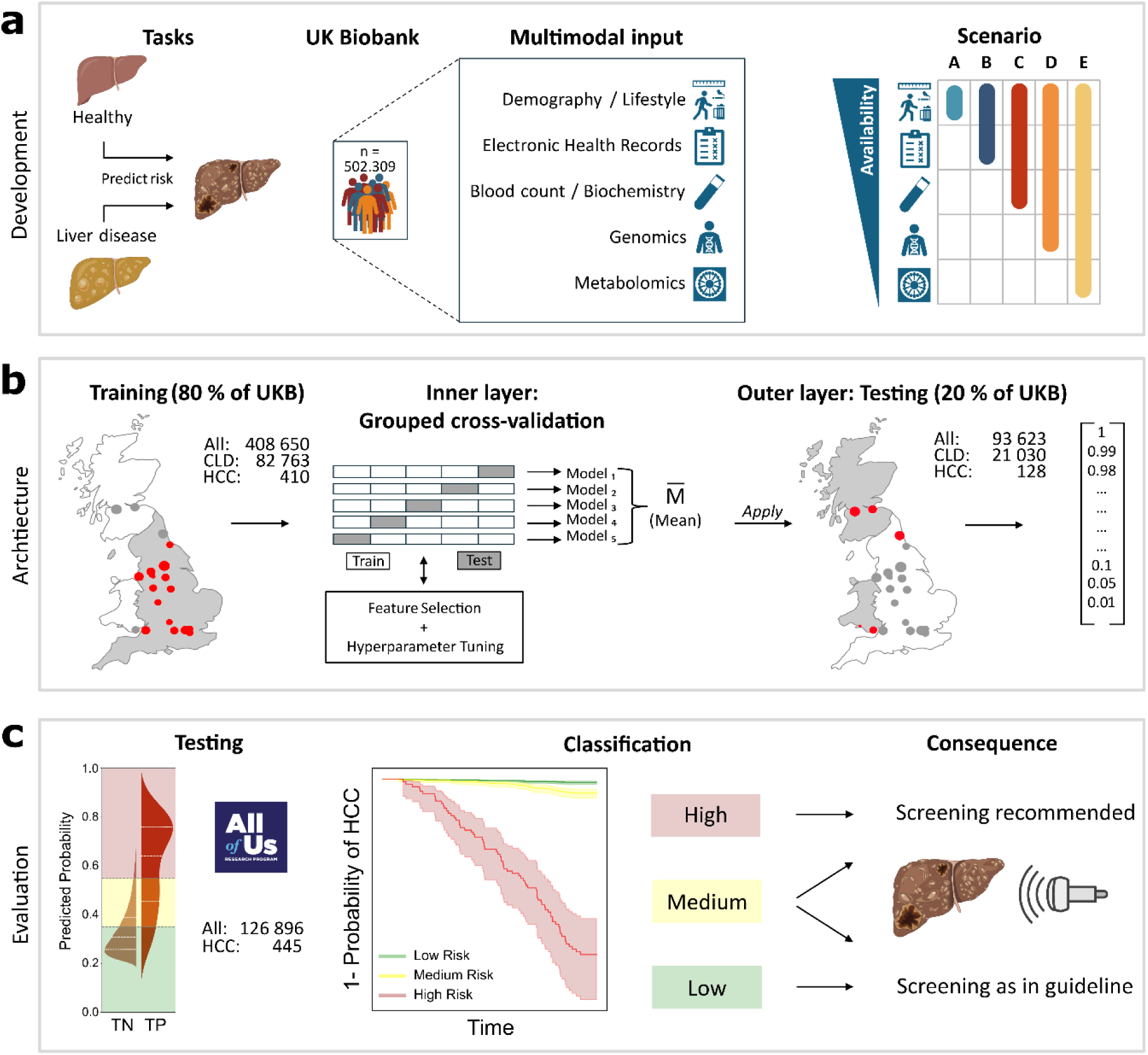
Study concept. **a** The task of predicting HCC occurrence was divided into prediction from a healthy cohort (“All”) and prediction among patients-at-risk (“PAR”). Multimodal data from UKB was extracted and scenarios set up according to availability of data on a patient’s trajectory in the healthcare system. **b** ML architecture with an inner-layer five-fold cross-validation, with a grouped-split approach, where each split (indicated by small squares) combines 4-5 assessment centers together,, with each split serving four times as training and once as validation set. Training data solely generated from UKB centers within England as indicated on the schematic map of Great Britain. The final model is a majority vote (M) built from the five generated ML models. M was then applied to the with-held test set built from UKB centers in Scotland/Wales (+Newcastle, for 80:20 balance between train/test) with numerical prediction output. **c** Evaluation and independent external testing of the model, including classification, time-to-event analysis and sub-group analysis. Thresholds are applied to rule-in for screening (when > “High”-threshold) or to rule-out for screening (when < “Low”-threshold).

We show that integration of multimodal data into ML models outperforms previous approaches for risk stratification of HCC development.

## Methods

### UK Biobank Cohort

The present investigation leverages data from the UKB, an expansive prospective cohort study initiated between 2006 and 2010. The UKB enrolled a diverse population of individuals aged 37-73 years, encompassing a total of 502,411 participants, with 502,309 participants currently available due to removal of consent of 102 participants. Initial evaluations were comprehensive, incorporating an array of anthropometric measurements, biospecimen collection, and the administration of multiple structured questionnaires. Ethical approval for the UKB study was granted by the Northwest Multi-Center Research Ethics Committee. Comprehensive methodological details, data accessibility, and acquisition protocols are publicly disclosed on the UKB’s official website (http://www.ukbiobank.ac.uk). Participants were ac-tively encouraged to partake in an inaugural clinical assessment, which was subsequently followed by longitudinal monitoring. All enrollees provided informed consent for genotypic analyses and the longitudinal linkage of their data to medical records. Periodic follow-up evaluations have been instituted to monitor alterations in health status and lifestyle variables.

### Primary survey data

Baseline characteristics were obtained through structured questionnaires. These self-reported measures included lifestyle factors such as alcohol consumption, smoking history, as well as medical history encompassing liver disease, diabetes, and obesity. Estimated values (Alcohol consumption per day; Pack years) were limited to the 99.9th percentile to minimize the influence of extreme outliers while preserving the overall distribution of the data. Physiologically plausible limits were set to further curate the dataset (Supplementary Table 3). Ethnicities in the “All Of Us Research Program” were inferred from questionnaires “Self-reported race” and “self-reported ethnicity”.

### Electronic health records

Electronic health records of UKB, documented by diagnosis codes at baseline assessment, were merged with EHR through ICD-Mapping (Supplementary Table 8), available via UKB through “hesin” and “hesin_diag”. Patients with an EHR entry of HCC before or in the first year after their first UKB visit were excluded from analysis (n=36) (Figure 2a) to prevent training on data of undetected HCC cases. Records were supplemented with UK death registers, with C220 documented here being treated equal to encoding via EHR. Training was performed on this data, while evaluation was restricted to all patients whose diagnosis of HCC was confirmed by data from the UK National Cancer Registry (UKB Resource 115558). EHR for a subset of 136 ICD codes (Supplementary Table 2, 3), consisting of a set of chronic liver diseases or signs of portal hypertension, cardiovascular risk factors, and gastrointestinal cancers except hepatobiliary cancers were included after performance of phenome wide association studies (Figure 2d). To prevent leakage of future diagnosis to the training data, only diagnosis codes up until 5 years after initial UKB visit were collected as features. EHR of “All Of Us” are curated from a variety of sources and then harmonized using the Observational Medical Outcomes Partnership (OMOP) Common Data Model to be stored in data dictionaries. Specific data relevant to our study were then extracted on their workbench platform using built-in cohort and dataset builders. Diagnostic instances of all relevant ICD codes for each individual were tabulated along with shifted dates, as any one individual may have the same diagnosis code recorded in their health record multiple times from multiple medical visits.

**Fig. 2:**
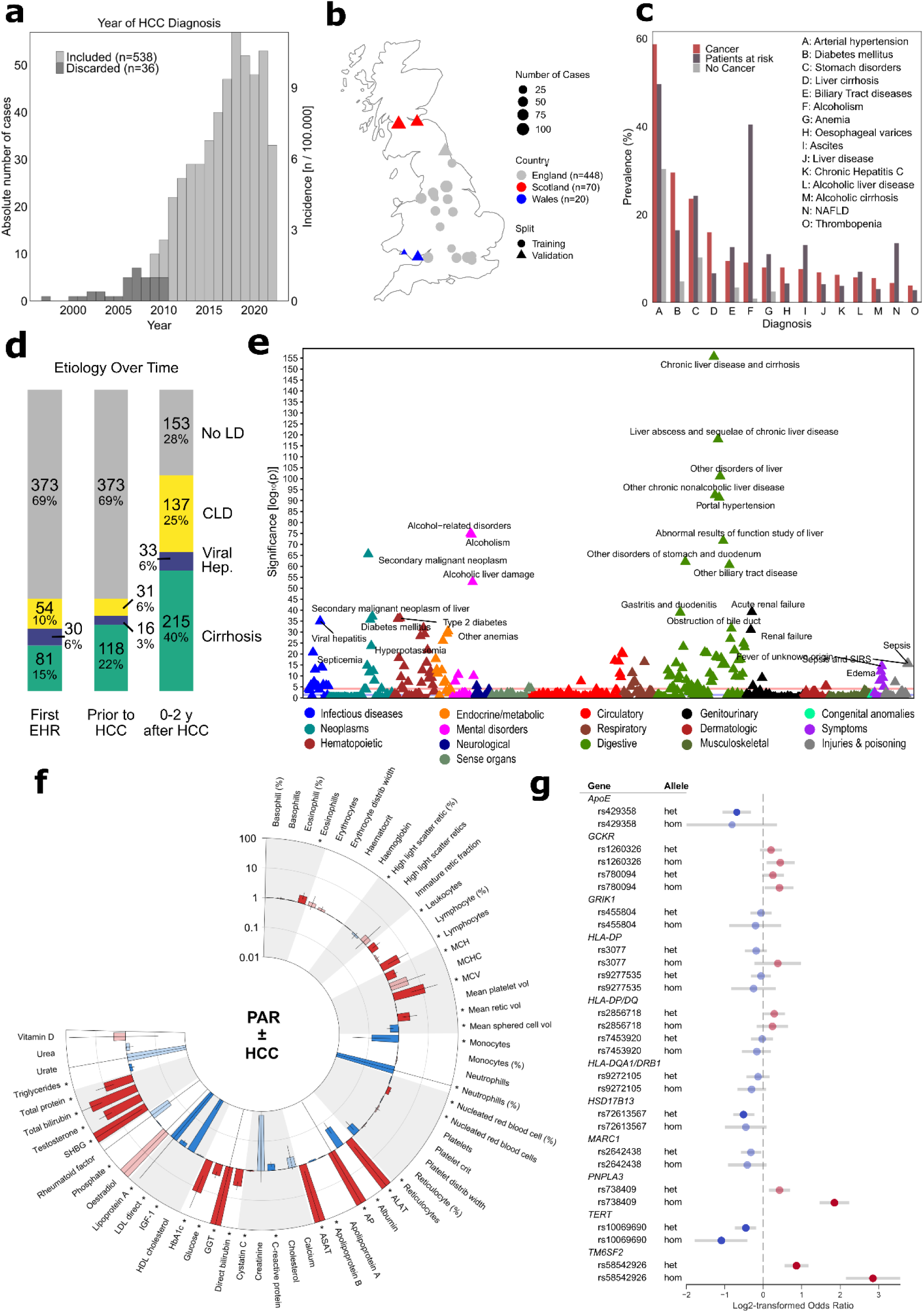
UK Biobank encompasses representable risk factors of HCC. **a** Histogram of first occurrences of HCC diagnosis in EHR in UKB. Cases discarded for analysis shown in dark gray (exclusion criteria see Methods). **b** Distribution of HCC cases in UK Biobank centers on a map of Great Britain, displayed by country association and mapping to training or validation cohort, mapped as number of cases per 500.000 **c** Prevalence of most common disease codes in EHR or self-reported for HCC cohort, patients-at-risk cohort and control group. Sorted by highest prevalence in the HCC group. **d** Ranking of etiologies of HCC cases (n=538) with total numbers + percentage. Bars indicate timepoints, with “First EHR” being the first-ever NHS-documented hospital stay, “Prior to HCC” the last stay before HCC diagnosis and “0-2y after HCC” indicating first diagnosis in the first two years after HCC diagnosis. No LD = No Liver Disease, CLD = Chronic Liver disease (except Cirrhosis or Viral Hepatitis) **e** Manhattan plot of phenome-wide-associations for individuals with HCC (C22.0) vs propensity-matched control population (on age, sex, bmi) (with P values (−log10). Highlighted are as-sociations with p values ≤ 0.05 (corrected for multiple testing by Bonferroni-correction. **f** Associations of blood biomarkers with HCC in CLD, displayed as Odds ratios (OR) and 95% confidence intervals for a one-standard deviation increase in each biomarker on the natural log scale, adjusted for age, sex and BMI. Significance defined as false discovery rate (FDR) controlled p < 0.01 in a linear model, marked with * and full saturation. Error bars limited to Y-limits for better readability. Mapping to UKB field IDs see Supplementary Table 5. **g** Associations of single-nucleotide polymorphisms with HCC occurrence displayed as log2-transformed Odds ratios + 95 % confidence interval, split between heterozygous (het) and homozygous (hom) occurrence, with color indicating direction of effect (blue = reduced risk, red = increased risk), and significance (Bonferroni-corrected p < 0.05) indicated by opacity, transparent=not-significant, opaque = significant.

### Blood count and biochemistry

Blood assays in UKB (Category 100080) were obtained at baseline assessment, with biological processing explained in detail on the UKB’s official website. Extracted features listed in Supplementary Table 1, 2, 3, 8. NaN-values were mean-imputed for all features except Oestradiol and Rheumatoid factor, which were excluded from analysis due to lack of data (Oestradiol 402.879 NAs; Rheumatoid factor 437.202 NAs). Cleaned features were min-max normalized (see “Data Processing”). Elevated liver enzymes were defined as follows, with sex-specific cut-offs for males/females, respectively: aspartate aminotransferase (50/35 U/L), alanine aminotransferase (50/35 U/L) and Gamma glutamyltransferase (60/40 U/L). Blood assays in “All Of us” are not assessed at a pre-defined baseline, but passed for every hospital visit of the participants. Values were harmonized to standardize units and then filtered to remove measurements made in an emergency or urgent care setting. Outliers more than four standard deviations from the mean were removed to filter out likely erroneous values, and then averaged over multiple instances to reduce the amount of missing data points.

### Genetic Data

Genetic profiling was conducted on a subset of 488,377 individuals within the UKB cohort, specifically targeting carriers of established hepatic risk variants after comprehensive literature research (Figure 2 f, Supplementary Table 7). Genotypes were downloaded using the UKB gfetch utility. Specific genotype calls for hepatic risk variants were extracted with PLINK, including alignment and quality control. For each single nucleotide polymorphism (SNP), a linear regression model corrected on age, sex and the first five principal components of the genotypes was performed. Our analysis stratified participants into non-carriers, heterozygous carriers, and homozygous carriers of the minor allele (refer to Supplementary Table 7 for details).

### Metabolomics

Utilizing nuclear magnetic resonance (NMR) spectroscopy, the UKB successfully characterized the metabolomic/lipidomic signatures of 248,266 participants within the cohort (Category 220). This high-throughput approach enabled the comprehensive quantification of a wide array of metabolites (n = 143), encompassing lipids, amino acids, and small-molecule intermediates, thereby providing an intricate biochemical snapshot pertinent to overall metabolic health. We extracted the 149 directly measured metabolomic features (see Supplementary Table 6), imputation and normalization procedures were performed in analogy to blood assays. All modeling analyses that included metabolomics data were performed only for the 248,266 participants for which metabolomics data was available.

### Data preprocessing UKB

Data extraction and preprocessing steps were performed with RStudio (Version 2023.12.1) and are publicly available in our GitHub repository (see “Code Availability”). Summary tables were created with the gtsummary package ^65^. Extracted data points are listed in Supplementary Tables. Data was processed separately for the following entities: Initial assessment (Demographics, Lifestyle, Self-reported diagnosis codes), Electronic Health Records, Blood/Serum, Single Nucleotide Polymorphisms, Metabolomics. Data was treated as independent factor level (categorical data) or imputed via mean imputation of continuous data. UKB Patients were stratified into six folds according to the assessment center (ID 54-0.0), with folds 1 to 5 as the basis for the five-fold cross-validation, and fold 6 as independent test set (Figure 2b, Supplementary Table 10). Outliers were limited to pre-defined physiological limits to allow for shared normalization between UKB and AOU to be applied (Supplementary Table 12).

### The All Of Us cohort

The All Of Us cohort is a longitudinal cohort with 409.420 participants as of 10/2024. The program collects and curates clinical, biological and environmental determinants of health and disease, with procedures described in detail elsewhere ^24^. Health data are obtained through electronic health records and participant surveys, which are available here: www.researchallofus.org/data-tools/survey-explorer/. AOU gathers EHR data from > 50 healthcare organizations, data stewards at provider organizations harmonize local data to the Observational Medical Outcomes Partnership (OMOP) Common Data Model, which can then be accessed by researchers. For details on biospecimen collection and pro-cessing, see https://allofus.nih.gov/funding-and-program-partners/biobank.

### Data preprocessing AOU

Preprocessing was performed in line with UKB preprocessing, with the following exceptions: Alcohol consumption was approximated by frequency and amount of drinks consumed with 14g Alcohol/drink as average. While UKB invites for a primary assessment, All Of Us is linked to hospital stays, therefore more prone to missing values. Participants with over 20 NA values were excluded from further analysis. AOU data was then converted to UKB units as documented in Supplementary Table 12.

### Computational infrastructure

Preprocessing and training of UKB data was performed on local workstations with Intel i7-i9 CPUs and 32 GB of RAM. Training times ranged between 10 and 30 minutes, without GPU requirement. Training times ranged from 10 minutes to a maximum of 2 hours for the biggest model. The cross-validations are parallelized to use all CPU-cores available to the user.

### Modeling and hyperparameter tuning

The modeling was conducted using scikit-learn version 1.2.2 ^42^. We chose the random forest classifier and extreme gradient boosting as default model architectures based on literature and practicability. We split the UKB-data into 2 sets, one for training incl. cross-validation with exclusively England-based UKB assessment centers and one for testing of the model performance (Glasgow, Edinburgh, Cardiff, Swansea, Newcastle). Training was carried out in a grouped five-fold cross-validation. Herein, each of the five folds (each consisting of 3-4 assessment centers, split see Supplementary Table 10-11), served for validation once and served for training in 4 out of 5 models. A grid search evaluating different hyperparameter combinations in the five-fold cross-validation revealed an optimal architecture with 50 estimators and a max depth of 3. This architecture was afterwards used for the training of all models to facilitate the comparison based on the input data and reduce the effect of different architectures. All five models derived from cross-validation were integrated into a majority-voting model, as an ensemble-classifier, the final model. This final model was then applied on the withheld test set (Held-out fold), as well as an external test dataset from the “All Of Us Cohort” for detailed performance and bias estimations.

### Interpretability

Feature importance was estimated as mean and standard deviation of accumulation of the impurity decrease within each tree via the scikit-learn library. SHapley Additive exPlanations (shap-library) were used for further interpretability with bootstrapping of individual participants feature importance, connecting optimal credit allocation with local explanations using the classic Shapley values from game theory and their related extensions ^53,66^.

### External Testing

External testing was performed in the research workbench of the “All Of Us Research Cohort” by an independent research group, with only access to the scripts, one hot encoder and model object. In-detail scripts and requirements available via our GitHub repository.

### Statistical analysis

Statistical comparisons between groups were performed using Welch’s two-sample t-tests for continuous variables and Chi-square tests for categorical variables. For categorical variables with low cell counts (<20), Chi-square approximation may be imprecise. Results are presented as mean ± standard deviation (SD) for continuous variables and absolute numbers (percentages) for categorical variables, with values < 20 masked for AOU to protect privacy as required by AOU. All statistical analyses were adjusted for multiple testing with false-discovery-rate or Bonferroni correction. For statistical comparison of AUROCs, two-sided DeLong tests were performed with Bonferroni post hoc correction. In depth documentation for all statistical analysis can be found in our GitHub repository. Statistical tests were calculated in R (Tables) or in Python (Models).

### Visualization

Visuals were created with RStudio 2024.04.0 as well as Python 3.9 matplotlib, scikit-learn, seaborn (0.12.2), shap ^53^ with all additional requirements being accessible via our GitHub repository. Figure assembly was performed with Inkscape 1.3.2, with integration of icons under common license from Microsoft, Flaticons, Bioicons, Healthicons.

## Results

### HCC cases in UK Biobank represent the general population

We hypothesized that population-based cohorts are well-suited to build HCC-prediction models. Firstly, they have a superior generalizability to the overall population compared to cohorts from tertiary centers and secondly, they uniquely provide rich data on phenotypic features even years before diagnosis of HCC ^19,22^. A total of 538 eligible HCC cases were observed in UKB (Supplementary Figure 1, Figure 2a) and 445 HCC cases in AOU, with a mean time to HCC diagnosis of 8.7 years, mean age at diagnosis of 70 ± 6.8 years in UKB and 62.1 ± 10.2 years in AOU (Supplementary Fig. 2a, b). Incidence was comparable to overall HCC incidence in the UK with 6-10 / 100,000 new cases per year (Figure 2a) ^25^, while more frequent in the AOU cohort (15-30 / 100.00, Figure 5a). All 22 centers of data acquisition showed similar abundance of HCC (Figure 2b, Supplementary Table 11). 399 of 538 HCC cases had a positive cancer-record in respective national cancer registries. Selection of 53 questionnaire categories on general and lifestyle information was based on the extensively characterized literature on known risk factors of HCC (Alcohol consumption, Smoking, Social status) ^26,27^ (Table 1, Supplementary Table 3). Selection of disease-codes for the prediction models was based on prior phenome-wide-association studies for HCC in UKB, controlled for age, sex, bmi and Townsend deprivation index. These revealed a multitude of significant correlations (Figure 2e, Supplementary Table 1, 2, 4, 8)), expectedly associating mostly liver diseases, but also alcoholism, diabetes and many more with risk of HCC. Notably, only 31 % of included HCC patients had a diagnosis of cirrhosis, viral hepatitis or other chronic liver diseases prior to HCC diagnosis, while 41 % were diagnosed with a liver disease in the two years after HCC diagnosis, and 28 % never received a diagnosis of liver disease via ICD10 (Figure 2 c, d, Supplementary Table 9, 13, 14). In addition to modeling the entire cohort (All), these pre-diagnosed patients were also investigated separately as a distinct “patients-at-risk” population (PAR). Next to previous diagnosis of steatotic liver diseases, viral hepatitis, or cirrhosis, the PAR population also encompassed cases with elevated liver enzymes at baseline examination (ICD-Codes in Supplementary Table 1, 2, 9). All blood count and serum parameters available diagnosis of HCC after controlling for age, sex, BMI and chronic liver disease (Figure 2f, Supplementary Table 5, 15, 16). Common single nucleotide polymorphisms known to increase risk of liver diseases such as MASLD, cirrhosis or HCC were included after confirmation via linear regression analysis on UKB data (Figure 2 g, Supplementary Table 7) ^6,28–35^. Nucleotide magnetic resonance spectroscopy metabolomics (NMR-Metabolomics) on 248,266 participants was investigated for possible correlating effects after correction for age, sex, bmi (Supplementary Fig. 2c, Supplementary Table 6). Of 143 metabolomic features, we observed 109 significant associations with HCC (Supplementary Table 17).

**Table 1:**
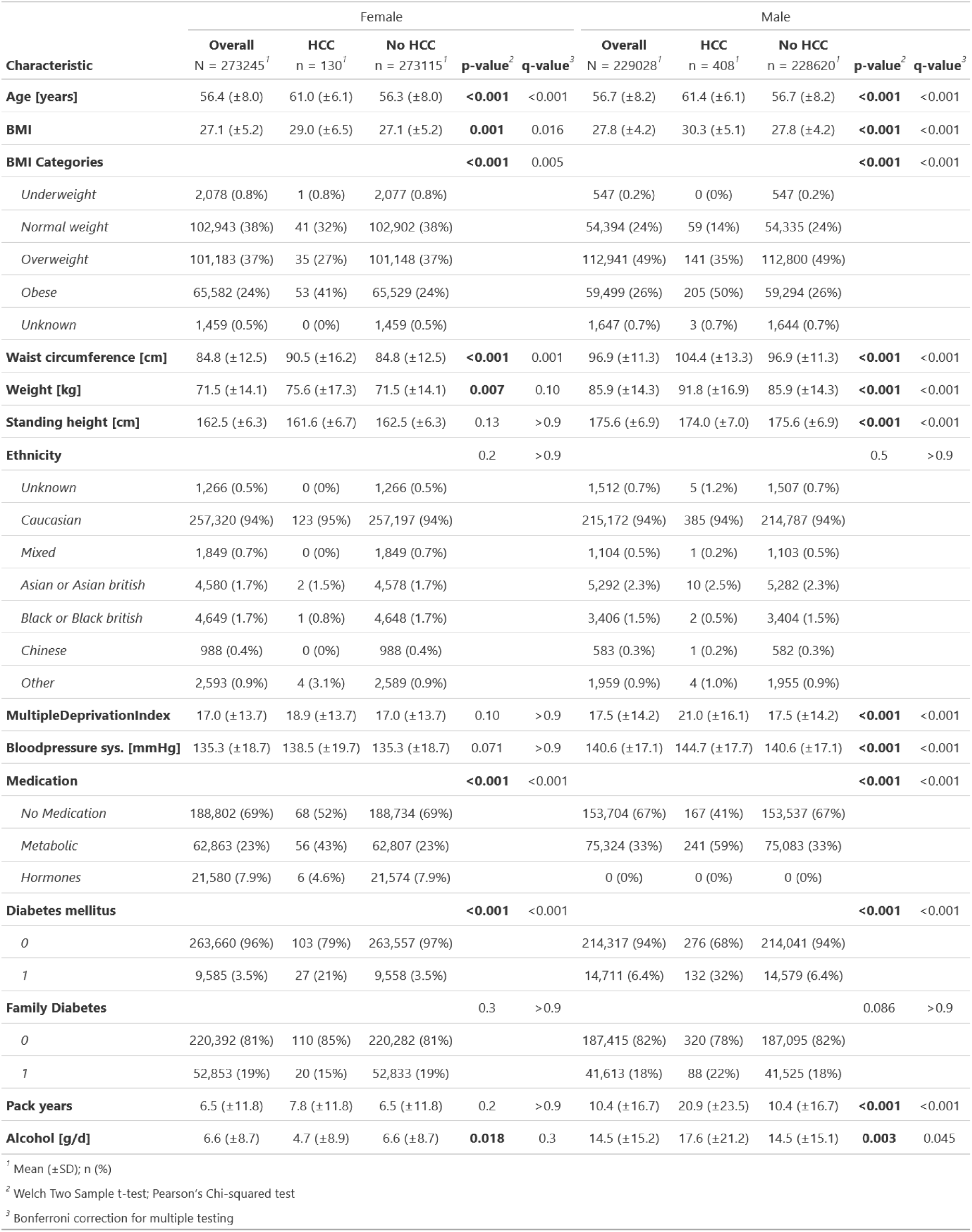
Baseline characteristics of the UK Biobank study population stratified by sex and HCC status. Baseline characteristics of study participants (N = 502 309) stratified by sex and hepatocellular carcinoma (HCC) status. Data are presented as mean (± SD) for continuous variables and n (%) for categorical variables. P-values were calculated using Student’s t-test for continuous variables and chi-square test for categorical variables. All continuous variables were assessed at time of UK Biobank enrollment. Q-values represent Bonferroni-adjusted p-values to account for multiple testing.

| Characteristic | Female |  |  |  |  | Male |  |  |  |  |
| --- | --- | --- | --- | --- | --- | --- | --- | --- | --- | --- |
|  | Overall<br>N = 273245<sup>1</sup> | HCC<br>n = 130<sup>1</sup> | No HCC<br>n = 273115<sup>1</sup> | p-value<sup>2</sup> | q-value<sup>3</sup> | Overall<br>N = 229028<sup>1</sup> | HCC<br>n = 408<sup>1</sup> | No HCC<br>n = 228620<sup>1</sup> | p-value<sup>2</sup> | q-value<sup>3</sup> |
| **Age [years]** | 56.4 (±8.0) | 61.0 (±6.1) | 56.3 (±8.0) | **<0.001** | <0.001 | 56.7 (±8.2) | 61.4 (±6.1) | 56.7 (±8.2) | **<0.001** | <0.001 |
| **BMI** | 27.1 (±5.2) | 29.0 (±6.5) | 27.1 (±5.2) | **0.001** | 0.016 | 27.8 (±4.2) | 30.3 (±5.1) | 27.8 (±4.2) | **<0.001** | <0.001 |
| **BMI Categories** |  |  |  | **<0.001** | 0.005 |  |  |  | **<0.001** | <0.001 |
| Underweight | 2,078 (0.8%) | 1 (0.8%) | 2,077 (0.8%) |  |  | 547 (0.2%) | 0 (0%) | 547 (0.2%) |  |  |
| Normal weight | 102,943 (38%) | 41 (32%) | 102,902 (38%) |  |  | 54,394 (24%) | 59 (14%) | 54,335 (24%) |  |  |
| Overweight | 101,183 (37%) | 35 (27%) | 101,148 (37%) |  |  | 112,941 (49%) | 141 (35%) | 112,800 (49%) |  |  |
| Obese | 65,582 (24%) | 53 (41%) | 65,529 (24%) |  |  | 59,499 (26%) | 205 (50%) | 59,294 (26%) |  |  |
| Unknown | 1,459 (0.5%) | 0 (0%) | 1,459 (0.5%) |  |  | 1,647 (0.7%) | 3 (0.7%) | 1,644 (0.7%) |  |  |
| **Waist circumference [cm]** | 84.8 (±12.5) | 90.5 (±16.2) | 84.8 (±12.5) | **<0.001** | 0.001 | 96.9 (±11.3) | 104.4 (±13.3) | 96.9 (±11.3) | **<0.001** | <0.001 |
| **Weight [kg]** | 71.5 (±14.1) | 75.6 (±17.3) | 71.5 (±14.1) | **0.007** | 0.10 | 85.9 (±14.3) | 91.8 (±16.9) | 85.9 (±14.3) | **<0.001** | <0.001 |
| **Standing height [cm]** | 162.5 (±6.3) | 161.6 (±6.7) | 162.5 (±6.3) | 0.13 | >0.9 | 175.6 (±6.9) | 174.0 (±7.0) | 175.6 (±6.9) | **<0.001** | <0.001 |
| **Ethnicity** |  |  |  | 0.2 | >0.9 |  |  |  | 0.5 | >0.9 |
| Unknown | 1,266 (0.5%) | 0 (0%) | 1,266 (0.5%) |  |  | 1,512 (0.7%) | 5 (1.2%) | 1,507 (0.7%) |  |  |
| Caucasian | 257,320 (94%) | 123 (95%) | 257,197 (94%) |  |  | 215,172 (94%) | 385 (94%) | 214,787 (94%) |  |  |
| Mixed | 1,849 (0.7%) | 0 (0%) | 1,849 (0.7%) |  |  | 1,104 (0.5%) | 1 (0.2%) | 1,103 (0.5%) |  |  |
| Asian or Asian british | 4,580 (1.7%) | 2 (1.5%) | 4,578 (1.7%) |  |  | 5,292 (2.3%) | 10 (2.5%) | 5,282 (2.3%) |  |  |
| Black or Black british | 4,649 (1.7%) | 1 (0.8%) | 4,648 (1.7%) |  |  | 3,406 (1.5%) | 2 (0.5%) | 3,404 (1.5%) |  |  |
| Chinese | 988 (0.4%) | 0 (0%) | 988 (0.4%) |  |  | 583 (0.3%) | 1 (0.2%) | 582 (0.3%) |  |  |
| Other | 2,593 (0.9%) | 4 (3.1%) | 2,589 (0.9%) |  |  | 1,959 (0.9%) | 4 (1.0%) | 1,955 (0.9%) |  |  |
| **MultipleDeprivationIndex** | 17.0 (±13.7) | 18.9 (±13.7) | 17.0 (±13.7) | 0.10 | >0.9 | 17.5 (±14.2) | 21.0 (±16.1) | 17.5 (±14.2) | **<0.001** | <0.001 |
| **Bloodpressure sys. [mmHg]** | 135.3 (±18.7) | 138.5 (±19.7) | 135.3 (±18.7) | 0.071 | >0.9 | 140.6 (±17.1) | 144.7 (±17.7) | 140.6 (±17.1) | **<0.001** | <0.001 |
| **Medication** |  |  |  | **<0.001** | <0.001 |  |  |  | **<0.001** | <0.001 |
| No Medication | 188,802 (69%) | 68 (52%) | 188,734 (69%) |  |  | 153,704 (67%) | 167 (41%) | 153,537 (67%) |  |  |
| Metabolic | 62,863 (23%) | 56 (43%) | 62,807 (23%) |  |  | 75,324 (33%) | 241 (59%) | 75,083 (33%) |  |  |
| Hormones | 21,580 (7.9%) | 6 (4.6%) | 21,574 (7.9%) |  |  | 0 (0%) | 0 (0%) | 0 (0%) |  |  |
| **Diabetes mellitus** |  |  |  | **<0.001** | <0.001 |  |  |  | **<0.001** | <0.001 |
| 0 | 263,660 (96%) | 103 (79%) | 263,557 (97%) |  |  | 214,317 (94%) | 276 (68%) | 214,041 (94%) |  |  |
| 1 | 9,585 (3.5%) | 27 (21%) | 9,558 (3.5%) |  |  | 14,711 (6.4%) | 132 (32%) | 14,579 (6.4%) |  |  |
| **Family Diabetes** |  |  |  | 0.3 | >0.9 |  |  |  | 0.086 | >0.9 |
| 0 | 220,392 (81%) | 110 (85%) | 220,282 (81%) |  |  | 187,415 (82%) | 320 (78%) | 187,095 (82%) |  |  |
| 1 | 52,853 (19%) | 20 (15%) | 52,833 (19%) |  |  | 41,613 (18%) | 88 (22%) | 41,525 (18%) |  |  |
| **Pack years** | 6.5 (±11.8) | 7.8 (±11.8) | 6.5 (±11.8) | 0.2 | >0.9 | 10.4 (±16.7) | 20.9 (±23.5) | 10.4 (±16.7) | **<0.001** | <0.001 |
| **Alcohol [g/d]** | 6.6 (±8.7) | 4.7 (±8.9) | 6.6 (±8.7) | **0.018** | 0.3 | 14.5 (±15.2) | 17.6 (±21.2) | 14.5 (±15.1) | **0.003** | 0.045 |

<sup>1</sup> Mean ( ± SD); n (%)
<sup>2</sup> Welch Two Sample t-test; Pearson's Chi-squared test
<sup>3</sup> Bonferroni correction for multiple testing

### Stepwise study architecture mimics data availability in real-world settings

We investigated predictive capacities of five different clinically relevant data modalities, namely demographics, electronic health records, bloodwork, genomics and metabolomics in two ways: first, for each modality individually, and second, using a stepwise approach reflecting the clinical availability of the modalities (Figure 1a). Predictive capacities were then assessed for both cohorts, All and PAR, resulting in a total of 20 model variations. Training and iterative hyperparameter tuning was performed in a five-fold cross-validation on data from only England-based centers of UKB, a total of 18 of 22 UKB centers. Testing of the final models was performed on untouched data of the 4 remaining centers, located either in Scotland or Wales (+ Newcastle) (n = 123 / 538) (Figure 1b, Supplementary Table 10, 11), as well as on the entire AOU cohort.

Compared to more complex deep learning techniques, decision-tree based machine learning models like random forest classifiers (RFC) have the advantage of direct explainability due to feature importance metrics, while being more data-efficient than neural networks or transformers ^36, 37^. We therefore included the decision-tree based models RFC and Extreme Gradient Boosting (XGB) in initial benchmarking experiments (Supplementary Fig. 3 a, b). Both models performed similarly, however RFC performance was more consistent, especially for models trained on reduced sample sizes. As RFC is also dominant in literature reports, we selected the RFC architecture as our baseline architecture to increase comparability. A grid-search for hyperparameter optimization on the internal cross-validation set revealed slim architectures (e.g. *max-depth = 3*, *n_estimators = 50*) to be beneficial despite the large size of the dataset (see Methods section).

### Machine learning is superior to linear risk scores for prediction of HCC in the general population

We hypothesized that accuracy of machine learning models would be superior to established liver-related risk assessment scores that are used to predict risk of HCC. We therefore performed a comparative analysis among available linear scores in the literature between aMAP ^38^, FIB-4 ^39^, APRI ^40^ and NFS ^41^ scores for prediction of HCC (Supplementary Fig. 3 c, d). Herein, the aMAP-score achieved the best performance of all linear scores with an AUROC of 0.79. We therefore used the aMAP-score as our benchmark throughout model performance evaluation. Evaluation on the withheld UKB test-set (90 HCC cases / 93533 controls for “All” and 72 cases / 20958 controls for “PAR”) revealed comparable performance throughout Model A-E for All and PAR (Figure 3 b, e, Table 3, Supplementary Fig. 3 e, Supplementary Table 18, 19), with blood parameters consistently as the most relevant modality (AUC 0.86 and AUC 0.87 for All/PAR respectively), followed by demographics (0.80 / 0.78), metabolomics (0.79 / 0.77), diagnosis (0.74 / 0.73) and SNPs (0.62 / 0.55). The single-modality models based on blood, demographics, and metabolomics all outperformed literature scores.

**Fig. 3:**
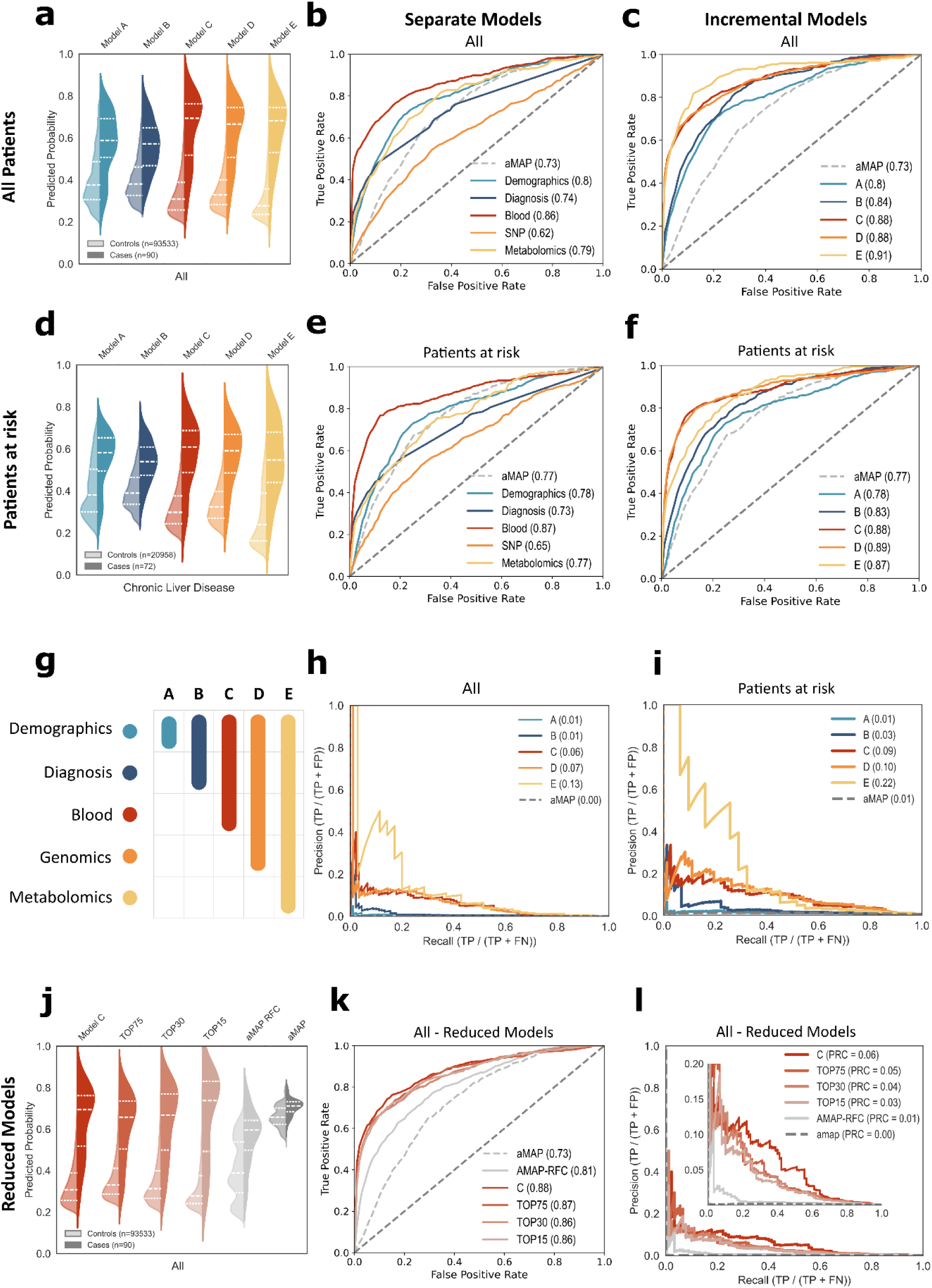
Prediction metrics. **a, d** Split violin plots displaying distribution of predicted probabilities for true negative (TN, left half violin) and true positive (TP, right half violin) cases per trained modality in test set for all patients (a) or PAR patients (d) in UKB test set. Quartiles (25th, 50th, 75th) displayed with dotted lines. **c, d, e, f** Model performance on the test set as receiver operating characteristic (ROC) curves and corresponding area under the curve (AUC) for either separate models (b, e) or combined scenarios A-E (c, f). Each line represents the performance of one majority vote model from five-fold cross-validation. Benchmark score ’aMAP’ (AGE + Male + (Albumin – Bilirubin) + Platelets²) and random guess depicted by dotted lines. **g** Legend for contribution of modalities per incremental model **h, i** Precision-recall curves for Model A-E, as well as aMAP score, with magnified section for precision = 0-0.2. TP = True Positives, FP = False Positives, FN= False Negatives. **j** Split violin distributions for prediction scores of models with reduced feature numbers (as in a/b) for UKB cohort. **k** ROC and indicated AUROC for reduced models in UKB, corresponding to j. **l** PRCs (as in h, i) for reduced models in UKB.

Clinical routine is multimodal, and a single modality likely never captures the true complexity. We therefore hypothesized a) that combination of modalities could improve model performance substantially, and b) that based on results from separate models, less affordable omics methods might not increase model performance substantially. Incremental models indeed revealed overall superior performance compared to separate models, with a plateau at an AU-ROC of 0.88 with model C, a combination of demographics, diagnosis and blood data (Fig 3 c, f, g). AUROCs could not be further improved by adding genetic information, and only improved by metabolomics for the All cohort, though this could not be tested externally.

Precision-recall curves (PRC) surprisingly revealed extremely low performance for the aMAP-score (Figure 3h, i) and other literature benchmarks (Supplementary Fig. 3f), with areas under the PRCs (AUPRC) of 0.00 to 0.01. The only exception for this was prediction of HCC based solely on the ICD-code Liver Cirrhosis (AUPRC of 0.14, Supplementary Fig. 3f), which proved superior AUPRCs also to ML models, while AUROCs for cirrhosis were worse (AUC 0.57 for UKB, 0.71 for AOU). For ML models, addition of blood data increased the AUPRC from negligible values to 0.06 (All) and 0.09 (PAR), with further increases both for D and E, although only for low-recall settings, i.e. very high risk cases. Meanwhile curves aligned completely for higher recall values, suggesting no additional contribution to overall discriminatory capabilities in a broader population.

### Explainability methods highlight known liver cancer risk factors

To increase practicability, we next performed an ablation study to reduce the number of features needed for the model (Fig. 3j-l). We employed a two-step approach, focusing firstly on routinely assessed features and secondly on removal of features with low relevance to the model, for which we extracted the models’ feature importance ^42^. First, a team of clinicians curated a set of 75 exclusively routine clinical features. For these, we then gradually removed features with lowest feature importance (TOP 75 features, TOP 30, TOP 15). Interestingly, a significant reduction in performance was observed between TOP75 and TOP30 (p = 0.0012, two-sided DeLong-Test), whereas performance of TOP30 and TOP15 did not differ significantly (p = 1, two-sided DeLong-Test). Mirroring this, a performance reduction was observed in AUPRCs from TOP75 to TOP30, but not from TOP30 to TOP15 (Figure 3l, Supplementary Table 23, 24). To corroborate that the models’ strong performances were due to the selected features and not merely the RFC architecture, we trained a RFC using only variables included in the aMAP score (AMAP-RFC). This model outperformed the original aMAP-score, while being significantly worse than the models with broader feature representation, suggesting that both the architecture and the feature amount contribute to the strong performance of our models.

We hypothesized that features relevant to the ML classifier would reflect the well characterized risk landscape also represented in linear risk scores. We therefore investigated the contribution weights of individual features for the model decision, and compared those per feature and per modality. We consistently observed the highest modality importance for blood-based features such as γ-GT, AST and ALT and platelets. Importantly however, feature importance was distributed more nuanced over the features, also incorporating measurements such as IGF-1, waist circumference and many more. In total, over 20 features contributed each with > 1 % (Figure 4 a, b, Supplementary Table 25). We further observed substantially different contributions to model prediction per modality. Blood data consistently had the highest contribution, both when assessed as mean or sum of feature importance per modality. This was followed by demography and lifestyle, EHR, and lastly genomics with negligible relevance for the model.

**Fig. 4:**
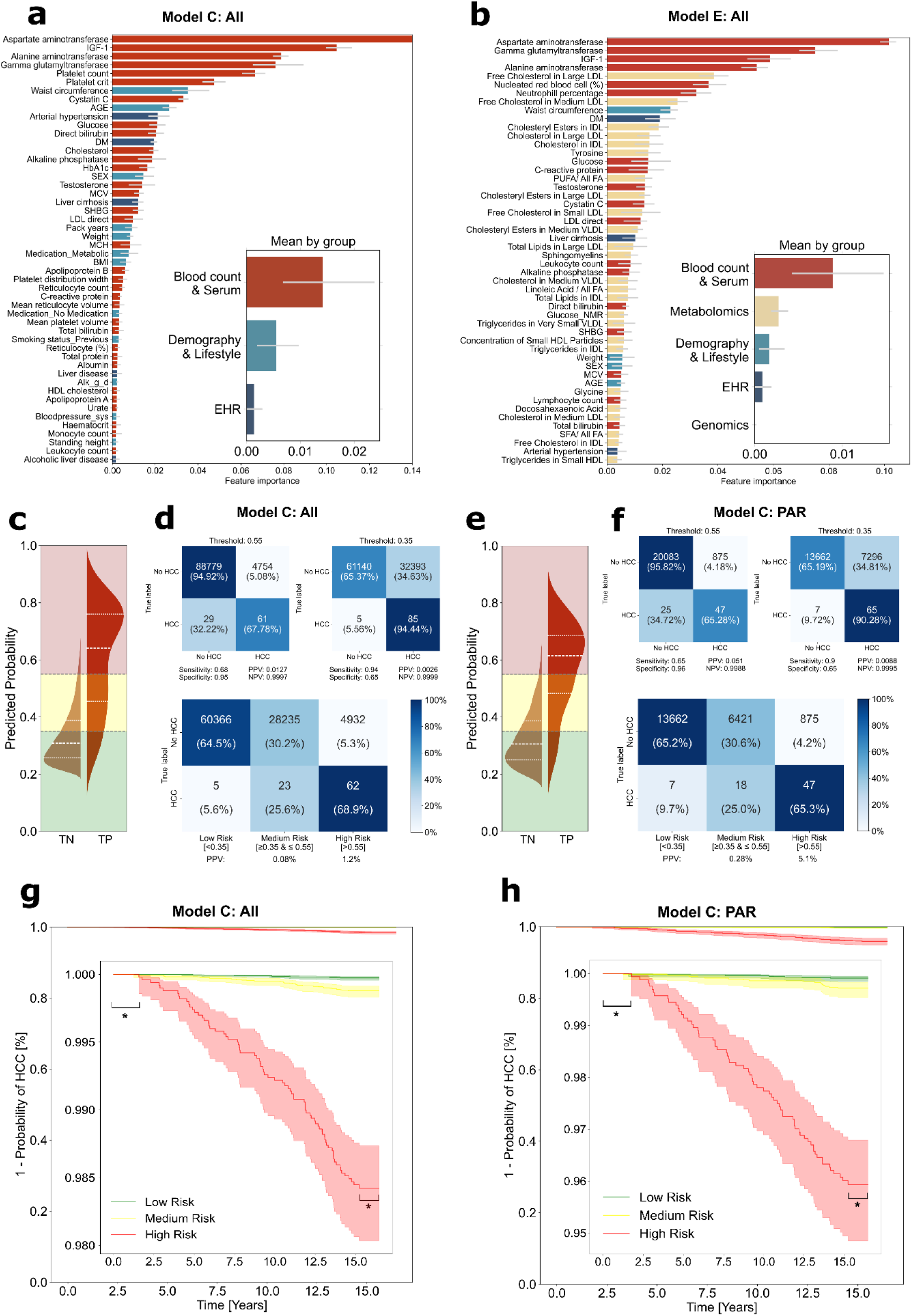
Interpretability of HCC risk prediction models. **a, b** Feature importances of the top-50 features as well as average by feature group (see Supplementary Material for corresponding features and groups) per indicated model, as mean ± SD of accumulation of the impurity decrease within each tree. **c, e** Split violin plots with displayed thresholding as low (green), middle (yellow) and high (red) risk groups, displaying distribution of predicted probabilities for true negatives (TN, left half violin) and true positive (TP, right half violin) cases per trained modality in test set for all patients (c) or PAR (e). Quartiles (25th, 50th, 75th) displayed with dotted lines. **d, f** Confusion matrices corresponding to predictions scores from c, e respectively, with indicated thresholds. Upper panels correspond to single thresholding, lower panels correspond to double thresholding. PPV = Positive predictive value. NPV = Negative predictive value. Coloring indicates relative percentage per class. **g**, **h** Kaplan-Meier curves for relative events per risk group over time. * indicates timeframes that were removed from analysis, either due to removal of events close to baseline examination or due to cut-off by 1st of January 2024.

### Thresholds create actionable classes for HCC risk estimation

Continuous scores are more accurate than classifications. However, in a clinical context they are often impractical, as they do not allow for clear decisions. A compromise between continuous scores and binary classification is commonly performed by categorizing predictions on an ordinal risk scale, e.g. in low-, middle-and high-risk groups, with a higher “rule-in” and a lower “rule-out” threshold. Based on the distribution of prediction values in the five-fold cross-validation, we established a three-class system, and evaluated the two thresholds on the with-held UKB test set, both for UKB “All” (Figure 4c, d) and “PAR” (Figure 4 e, f). In both cohorts, > 65 % of HCC cases could hereby be classified in the high-risk group with a positive predictive value (PPV) of 1.2 % (“All”) and 5.1 % (“PAR”), negative predictive values (NPV) of 0.9999 (“All”) and 0.9995 (“PAR”). High-and medium-risk groups together accounted for 94.5 % (“All”) and 90.3 % (“PAR”) of HCC cases. Still, as a low-incidence disease, this performance is challenged by a high number of false positive cases. We conducted a comprehensive comparison between the true positive (TP) and false negative (FN) groups, as well as true negative (TN) and false positive (FP) groups. Firstly, there was no significant difference between the time-to-cancer for TP (9.6 ± 3,7 years) compared to FN 9 ± 3,8 years). Notably, for UKB, we found a significantly higher incidence of FN among females compared to males (Table 3). Furthermore, distinct patterns emerged among the false negative group, characterized by a lower prevalence of obesity, smoking, and alcohol consumption.

### Generalizable models for HCC risk stratification

Finally, we hypothesized that our models would be generalizable to an independent population. We therefore used all participants of the “All Of Us Research Program” with at least one instance of available blood data and baseline demographic information (n = 126 896, 445 cases of HCC). Incidence of HCC (Figure 5a) and age distribution at first diagnosis (Supplementary Fig. 2b) was comparable to UKB data and to literature reports ^43^. Ethnicity distribution among controls and cases was much more diverse than in the UKB (Figure 5b, Table 2), and we observed a clear tendency towards better EHR representation among HCC cases, especially for liver cirrhosis (Figure 5c, Table 2). After rigorous preprocessing, the models TOP75, TOP30, TOP15 and AMAP-RFC were applied to all AOU patients along linear risk scores. All ML models achieved high AUROCs, interestingly without performance loss for models with reduced features, with also AUPRCs comparable to AUPRCs from the UKB All test cohort (Figure 5, Supplementary Table 20, 21). Again, ML-models consistently outperformed linear risk models (Figure 5 d, e, f, Supplementary Fig. 3c, d, f), despite linear risk models performing slightly better in AOU than in the UKB cohort. While the relative distribution of prediction values for controls and cases was very similar to UKB, we observed statistically significant higher overall prediction values in AOU (0.38 ± 0.15 (AOU) vs 0.33 ± 0.13 (UKB) for controls, p < 0.0001, and 0.68 ± 0.19 (AOU) vs 0.62 ± 0.22 (UKB) for HCC cases, p = 0.008, for Model TOP15, see Supplementary Table 22), reflecting the overall differences in disease burden between the two populations.

**Fig. 5:**
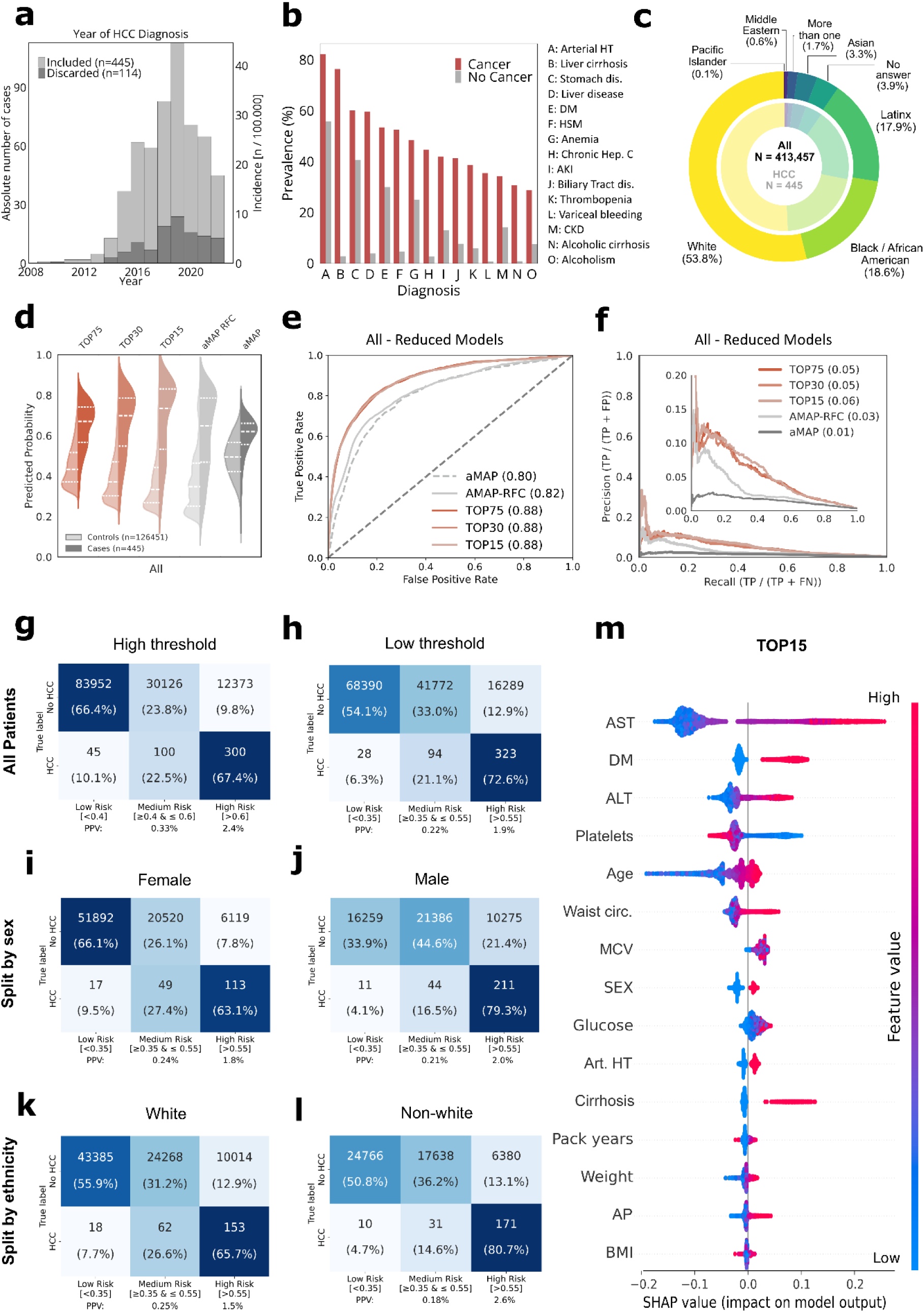
Machine learning models are generalizable to independent populations. **a** Histogram of first occurrences of HCC diagnosis in EHR in All Of Us. b Prevalence of the most common disease codes for HCC group (red) and control group (gray), sorted by highest prevalence in HCC group. Arterial HT = Arterial Hypertension, Dis.= Disease, DM = Diabetes mellitus, HSM = Hepatosplenomegaly, AKI = Acute kidney injury, CKD = chronic kidney disease. **c** Ethnic distribution of all participants (outer circle) and HCC cases (inner circle) in All Of Us, corresponding to self-reported survey categories. **c** Ranking of etiologies of HCC cases (n=445) with total numbers + percentage of etiologies in All Of Us. **d** Split violin plots displaying distribution of predicted probabilities for true negative (TN, left half violin) and true positive (TP, right half violin) cases per trained modality in All Of Us. **e** Model performance on All Of us as ROC curves and corresponding AUCs. **f** Precision-recall curves corresponding to models from d, e. **g-l** Confusion matrices corresponding to prediction scores from d-f, split by high vs lower thresholds, female vs male sex or by ethnicity (white vs non-white). **m** Shapley feature importance analysis for Model TOP15, with each datapoint representing a single participant. AST = Aspartate aminotransferase, DM = Diabetes mellitus, ALT = Alanine aminotransferase, HT = hypertension, AP = Alkaline phosphatase. Distance from 0.0 on x-axis indicates feature importance, color indicates direction of feature.

**Table 2:** Baseline characteristics of All of Us Research Program study population stratified by sex and HCC status. Baseline characteristics of study participants (N = 126 896) stratified by sex and hepatocellular carcinoma (HCC) status. Data are presented as mean (± SD) for continuous variables and n (%) for categorical variables. P-values were calculated using Student’s t-test for continuous variables and chi-square test for categorical variables. Fea-tures with less than 20 individual data points grouped together are masked (displayed as > 20) as is the requirement of the All Of Us Research Workbench. Q-values represent Bonferroni-adjusted p-values to account for multiple testing.

| Characteristic | Female |  |  |  |  | Male |  |  |  |  |
| --- | --- | --- | --- | --- | --- | --- | --- | --- | --- | --- |
|  | Overall<br>N = 78710<sup>1</sup> | HCC<br>n = 179<sup>1</sup> | No HCC<br>n = 78531<sup>1</sup> | p-value<sup>2</sup> | q-value<sup>2</sup> | Overall<br>N = 48186<sup>1</sup> | HCC<br>n = 266<sup>1</sup> | No HCC<br>n = 47920<sup>1</sup> | p-value<sup>2</sup> | q-value<sup>2</sup> |
| **Age [years]** | 54.0 (±16.2) | 61.1 (±11.9) | 54.0 (±16.2) | **<0.001** | <0.001 | 59.1 (±15.6) | 63.9 (±8.8) | 59.0 (±15.6) | **<0.001** | <0.001 |
| **BMI** | 30.8 (±8.3) | 30.5 (±7.3) | 30.8 (±8.3) | 0.6 | >0.9 | 29.7 (±6.6) | 28.8 (±5.2) | 29.7 (±6.6) | **0.006** | 0.074 |
| **Waist circumference [cm]** | 95.8 (±17.3) | 98.2 (±14.8) | 95.8 (±17.3) | **0.031** | 0.4 | 102.1 (±15.5) | 102.3 (±12.7) | 102.1 (±15.5) | 0.8 | >0.9 |
| **Weight [kg]** | 81.3 (±22.8) | 79.8 (±21.7) | 81.3 (±22.8) | 0.4 | >0.9 | 91.9 (±22.0) | 86.8 (±18.1) | 91.9 (±22.0) | **<0.001** | <0.001 |
| **Standing height [cm]** | 162.3 (±7.1) | 161.3 (±7.1) | 162.3 (±7.1) | **0.046** | 0.6 | 175.7 (±7.8) | 173.3 (±7.8) | 175.7 (±7.8) | **<0.001** | <0.001 |
| **Self-reported ethnicity** |  |  |  | **0.019** | 0.2 |  |  |  | **0.002** | 0.029 |
| Asian | 1,779 (2.3%) | <20 | 1,777 (2.3%) |  |  | 1,124 (2.3%) | <20 | 1,111 (2.3%) |  |  |
| Black / African American | 13,505 (17%) | 40 (22%) | 13,465 (17%) |  |  | 6,924 (14%) | 42 (16%) | 6,882 (14%) |  |  |
| Latinx | 14,532 (18%) | 43 (24%) | 14,489 (18%) |  |  | 6,505 (13%) | 51 (19%) | 6,454 (13%) |  |  |
| Middle Eastern | 334 (0.4%) | <20 | 333 (0.4%) |  |  | 318 (0.7%) | <20 | 316 (0.7%) |  |  |
| More than one | 1,183 (1.5%) | <20 | 1,183 (1.5%) |  |  | 541 (1.1%) | <20 | 536 (1.1%) |  |  |
| No answer | 2,132 (2.7%) | <20 | 2,123 (2.7%) |  |  | 1,549 (3.2%) | <20 | 1,539 (3.2%) |  |  |
| Pacific Islander | 59 (<0.1%) | <20 | 59 (<0.1%) |  |  | 64 (0.1%) | <20 | 63 (0.1%) |  |  |
| White | 45,186 (57%) | 84 (47%) | 45,102 (57%) |  |  | 31,161 (65%) | 142 (53%) | 31,019 (65%) |  |  |
| **MultipleDeprivationIndex** | 0.3 (±0.1) | 0.3 (±0.1) | 0.3 (±0.1) | **<0.001** | <0.001 | 0.3 (±0.1) | 0.3 (±0.1) | 0.3 (±0.1) | **<0.001** | 0.002 |
| **Bloodpressure sys. [mmHg]** | 125.0 (±17.4) | 129.2 (±19.3) | 124.9 (±17.4) | **0.003** | 0.045 | 130.2 (±16.6) | 130.7 (±16.7) | 130.2 (±16.6) | 0.7 | >0.9 |
| **Medication** |  |  |  | 0.7 | >0.9 |  |  |  | 0.10 | >0.9 |
| Hormones | 849 (1.1%) | <20 | 848 (1.1%) |  |  | 320 (0.7%) | <20 | 317 (0.7%) |  |  |
| Metabolic | 15,545 (20%) | 38 (21%) | 15,507 (20%) |  |  | 11,939 (25%) | 52 (20%) | 11,887 (25%) |  |  |
| No Medication | 62,316 (79%) | 140 (78%) | 62,176 (79%) |  |  | 35,927 (75%) | 211 (79%) | 35,716 (75%) |  |  |
| **Diabetes mellitus** | 21,829 (28%) | 99 (55%) | 21,730 (28%) | **<0.001** | <0.001 | 16,285 (34%) | 139 (52%) | 16,146 (34%) | **<0.001** | <0.001 |
| **Family Diabetes** | 10,980 (14%) | <20 | 10,964 (14%) | 0.067 | 0.9 | 4,442 (9.2%) | <20 | 4,425 (9.2%) | 0.14 | >0.9 |
| **Pack years** | 4.8 (±11.7) | 8.6 (±18.7) | 4.8 (±11.7) | **0.007** | 0.094 | 8.7 (±17.1) | 10.3 (±16.7) | 8.7 (±17.1) | 0.11 | >0.9 |
| **Alcohol [g/d]** | 3.4 (±7.2) | 1.7 (±6.1) | 3.4 (±7.2) | **<0.001** | 0.003 | 6.2 (±11.7) | 5.6 (±14.2) | 6.2 (±11.7) | 0.4 | >0.9 |

<sup>1</sup> Mean ( ± SD); n (%)
<sup>2</sup> Welch Two Sample t-test; Pearson's Chi-squared test
<sup>3</sup> Bonferroni correction for multiple testing

Again, model performance was better for male than for female cases, although with less pronounced differences (Figure 5, Table 3). Interestingly, model performance for European vs non-European populations did not differ significantly and did especially not lean towards better performance for the European population, despite training on 94 % data from “Caucasian” ethnicity, representing the distribution in UKB (Figure 5, Supplementary Table 21).

**Table 3:** Confusion matrix sub-analysis for both UKB and AOU cohort. Baseline characteristics split by bins of confusion matrix (FN = False negatives, TP = True positives, FP = False positives, TN = True negatives) for both UKB and AOU cohorts at the “rule-in” threshold of 0.55, with > 0.55 = High Risk / positive class, < 0.55 = Low or middle risk / negative class. Categories for ethnicity were unified and simplified to allow for shared visualization. Features with less than 20 individual data points grouped together are masked (displayed as > 20) as is the requirement of the All Of Us Research Workbench. For logical features (True/False), only positive class is displayed. Percentages represent a fraction of population per column. Only categorical features were subjected to comparison of significant differences. q-values represent Bonferroni-adjusted p-values to account for multiple testing.

| Characteristic | UKB |  |  |  |  |  |  | AOU |  |  |  |  |  |  |
| --- | --- | --- | --- | --- | --- | --- | --- | --- | --- | --- | --- | --- | --- | --- |
|  | Overall<br>N = 93623<sup>1</sup> | FN<br>n = 26<sup>1</sup> | TP<br>n = 64<sup>1</sup> | FP<br>N = 7,887<sup>1</sup> | TN<br>N = 85,646<sup>1</sup> | p-value<sup>2</sup> | q-value<sup>3</sup> | Overall<br>N = 126896<sup>1</sup> | FN<br>n = 121<sup>1</sup> | TP<br>n = 324<sup>1</sup> | FP<br>N = 16,394<sup>1</sup> | TN<br>N = 110,057<sup>1</sup> | p-value<sup>2</sup> | q-value<sup>3</sup> |
| Prediction Score | 0.3 (±0.1) | 0.4 (±0.1) | 0.8 (±0.1) | 0.7 (±0.1) | 0.3 (±0.1) |  |  | 0.4 (±0.1) | 0.4 (±0.1) | 0.8 (±0.1) | 0.7 (±0.1) | 0.3 (±0.1) |  |  |
| Age [years] | 56.5 (±8.0) | 60.2 (±6.5) | 61.6 (±5.8) | 59.5 (±6.3) | 56.2 (±8.1) |  |  | 55.9 (±16.2) | 63.0 (±13.9) | 62.7 (±8.5) | 60.0 (±13.4) | 55.3 (±16.5) |  |  |
| SEX |  |  |  |  |  | <0.001 | <0.001 |  |  |  |  |  | <0.001 | <0.001 |
| Female | 51,433 (55%) | 11 (42%) | 8 (13%) | 2,277 (29%) | 49,137 (57%) |  |  | 78,710 (62%) | 66 (55%) | 113 (35%) | 6,119 (37%) | 72,412 (66%) |  |  |
| Male | 42,190 (45%) | 15 (58%) | 56 (88%) | 5,610 (71%) | 36,509 (43%) |  |  | 48,186 (38%) | 55 (45%) | 211 (65%) | 10,275 (63%) | 37,645 (34%) |  |  |
| Ethnicity |  |  |  |  |  | 0.3 | >0.9 |  |  |  |  |  | <0.001 | <0.001 |
| Asian | 790 (0.8%) | 0 (0%) | 1 (1.6%) | 55 (0.7%) | 734 (0.9%) |  |  | 2,903 (2.3%) | <20 | <20 | 267 (1.6%) | 2,621 (2.4%) |  |  |
| Black | 207 (0.2%) | 0 (0%) | 0 (0%) | 10 (0.1%) | 197 (0.2%) |  |  | 20,429 (16%) | <20 | 65 (20%) | 2,618 (16%) | 17,729 (16%) |  |  |
| Caucasian | 91,653 (98%) | 26 (100%) | 63 (98%) | 7,755 (98%) | 83,809 (98%) |  |  | 76,347 (60%) | 80 (66%) | 146 (45%) | 9,825 (60%) | 66,296 (60%) |  |  |
| Latinx | 0 (0%) | 0 (0%) | 0 (0%) | 0 (0%) | 0 (0%) |  |  | 21,037 (17%) | <20 | 79 (24%) | 2,919 (18%) | 18,024 (16%) |  |  |
| Other/Unknown | 973 (1.0%) | 0 (0%) | 0 (0%) | 67 (0.8%) | 906 (1.1%) |  |  | 6,180 (4.9%) | <20 | 23 (7.1%) | 765 (4.7%) | 5,387 (4.9%) |  |  |
| BMI | 27.7 (±4.8) | 30.0 (±7.6) | 30.3 (±4.3) | 30.3 (±5.2) | 27.4 (±4.7) |  |  | 30.4 (±7.7) | 28.5 (±5.9) | 29.9 (±6.3) | 31.2 (±7.5) | 30.3 (±7.7) |  |  |
| Waist circumference [cm] | 90.3 (±13.3) | 98.2 (±18.7) | 102.6 (±12.5) | 100.2 (±13.5) | 89.4 (±12.9) |  |  | 98.2 (±16.9) | 97.4 (±14.0) | 101.9 (±13.4) | 103.2 (±17.0) | 97.4 (±16.8) |  |  |
| Weight [kg] | 78.1 (±15.9) | 86.5 (±21.2) | 89.4 (±16.2) | 88.3 (±17.3) | 77.1 (±15.4) |  |  | 85.3 (±23.1) | 80.0 (±18.3) | 85.5 (±20.3) | 90.6 (±23.8) | 84.5 (±22.9) |  |  |
| Standing height [cm] | 167.7 (±9.2) | 170.4 (±9.7) | 171.3 (±8.0) | 170.6 (±9.0) | 167.5 (±9.2) |  |  | 167.4 (±9.8) | 167.3 (±9.6) | 168.9 (±9.5) | 170.3 (±10.1) | 167.0 (±9.7) |  |  |
| MultipleDeprivationIndex | 16.4 (±14.2) | 18.0 (±11.0) | 23.6 (±19.3) | 17.9 (±15.0) | 16.2 (±14.1) |  |  | 0.3 (±0.1) | 0.3 (±0.1) | 0.3 (±0.1) | 0.3 (±0.1) | 0.3 (±0.1) |  |  |
| Bloodpressure sys. [mmHg] | 138.4 (±16.6) | 141.4 (±13.5) | 146.8 (±18.7) | 143.4 (±17.0) | 138.0 (±16.5) |  |  | 126.9 (±17.3) | 129.3 (±18.6) | 130.4 (±17.5) | 129.6 (±17.8) | 126.5 (±17.2) |  |  |
| Medication |  |  |  |  |  | <0.001 | <0.001 |  |  |  |  |  | <0.001 | <0.001 |
| Hormones | 3,791 (4.0%) | 0 (0%) | 0 (0%) | 67 (0.8%) | 3,724 (4.3%) |  |  | 1,169 (0.9%) | <20 | <20 | 140 (0.9%) | 1,025 (0.9%) |  |  |
| Metabolic | 26,245 (28%) | 15 (58%) | 32 (50%) | 4,107 (52%) | 22,091 (26%) |  |  | 27,484 (22%) | 34 (28%) | 56 (17%) | 4,131 (25%) | 23,263 (21%) |  |  |
| No Medication | 63,587 (68%) | 11 (42%) | 32 (50%) | 3,713 (47%) | 59,831 (70%) |  |  | 98,243 (77%) | 86 (71%) | 265 (82%) | 12,123 (74%) | 85,769 (78%) |  |  |
| Diabetes mellitus | 4,086 (4.4%) | 2 (7.7%) | 19 (30%) | 1,176 (15%) | 2,889 (3.4%) | <0.001 | <0.001 | 38,114 (30%) | 48 (40%) | 190 (59%) | 8,179 (50%) | 29,697 (27%) | <0.001 | <0.001 |
| Family Diabetes | 16,785 (18%) | 8 (31%) | 17 (27%) | 1,649 (21%) | 15,111 (18%) | <0.001 | <0.001 | 15,422 (12%) | <20 | 22 (6.8%) | 1,811 (11%) | 13,578 (12%) | <0.001 | <0.001 |
| Pack years | 8.8 (±15.4) | 18.3 (±27.3) | 25.0 (±26.8) | 13.2 (±19.7) | 8.4 (±14.8) |  |  | 6.3 (±14.1) | 7.8 (±14.5) | 10.3 (±18.5) | 9.2 (±17.2) | 5.8 (±13.5) |  |  |
| Alcohol [g/d] | 10.2 (±12.8) | 10.2 (±13.4) | 18.6 (±21.4) | 15.0 (±17.8) | 9.7 (±12.1) |  |  | 4.5 (±9.3) | 2.3 (±6.2) | 4.7 (±13.2) | 6.4 (±13.6) | 4.2 (±8.4) |  |  |
| Liver cirrhosis | 171 (0.2%) | 1 (3.8%) | 18 (28%) | 83 (1.1%) | 69 (<0.1%) | <0.001 | <0.001 | 3,573 (2.8%) | 51 (42%) | 289 (89%) | 2,284 (14%) | 949 (0.9%) | <0.001 | <0.001 |

<sup>1</sup> Mean (±SD); n (%) <sup>2</sup> Pearson's Chi-squared test <sup>3</sup> Bonferroni correction for multile testing
<sup>1</sup> Mean (±SD); n (%)
<sup>2</sup> Pearson's Chi-squared test
<sup>3</sup> Bonferroni correction for multiple testing

## Discussion

Our study shows that simple, interpretable machine learning algorithms can accurately stratify patients according to their individual risk of developing HCC on a population scale. We present these models along with systematic evaluation of risk scores proposed in literature. Evaluation on two of the largest available cohorts worldwide shows superior performance of the developed ML models compared to all publicly available risk scores and provide robust explainability for our algorithms.

Current HCC screening algorithms incorporate cirrhosis, a highly specific, but insensitive risk factor for HCC, as the sole inclusion criterion for apparative screening. This is problematic as firstly, compensated cirrhosis can be occult and therefore not diagnosed in a large fraction of patients and secondly, the percentage of HCC in non-cirrhotic patients is steadily growing, widening the population at risk. This gap in screening facilitates late-stage diagnosis and high mortality of HCC. Non-invasive risk scores used as surrogates for liver fibrosis and predictors of liver-related mortality are usually developed in small, tertiary-hospital-based cohorts, lack accuracy, and do not represent the complex individual risk landscape ranging from diagnosis, lifestyle, demographics to bloodwork. Further, they are rarely validated on population-level. We hypothesized that population-based cohorts with longitudinal follow-up are the ideal foundation for generalizable risk stratification in HCC. This allowed for simulation of a prospective evaluation on two large, independent test sets, according to the “Transparent reporting of a multivariable prediction model for individual prognosis or diagnosis (TRIPOD)”, with firstly, independent testing on all data from UKB centers in Scotland and Wales (Nonrandom split-sample development, TRIPOD 2b), and secondly, the entire “All Of Us” cohort (Separate test data set, TRIPOD 3) ^44^.

To account for distinct pre-test probabilities, especially between primary and tertiary-care centers, we evaluated all our models in a general population and for a subset of patients-at-risk, with pre-diagnosed liver disease or elevated liver enzymes at baseline. We show that, based on performance in two distinct populations, the aMAP-score is the best-performing publicly available linear risk score for HCC. However, despite a modest accuracy, negligible precision and recall lead to an extremely high number of false positive cases for HCC risk. The observed imbalance between metrics is presumed to be caused by the inherent properties of receiver-operating-characteristics. These can cause overestimation of model performance in imbalanced data, which is the case in virtually every screening scenario for low-prevalence diseases ^45^ ^46,47^. In contrast to aMAP, presence of cirrhosis prior to HCC was a highly specific, but insensitive predictor of HCC. This suggests that cirrhosis alone is a good “rule-in” discriminator for HCC-screening, but an insufficient “rule-out” discriminator in a broad population, while it is still applied here as a sole means of decision-support in various guidelines ^7,48^.

Our study presents the first systematic comparisons for relevance of independent data modalities. We show that training machine learning models on standard blood values like transaminases and platelets together with EHR data and lifestyle is sufficient for accurate risk stratification for HCC, in line with previous work for other cancer types and relevant variables in linear risk scores for HCC ^49,50,38–40^. Interestingly, genomic data did not increase model performance despite representable prevalences of known genetic risk factors like PNPLA3 ^6^ (Figure 2g). In line with previous reports, this questions the relevance of polygenic risk scores (PRS) for risk prediction of HCC compared to phenotypic modeling ^8, 51,52^. Meanwhile, metabolomics did increase model performance to some extent, but they are not routinely measured, more expensive and eventually not necessary given the satisfactory performance of models based on routine clinical data alone.

Investigation of model explainability highlights our models, especially the slimmer models TOP15 -TOP75, as clinically feasible and interpretable tools ^53^. We confirm liver enzymes, platelets, demographic features such as age and weight, but also arterial hypertension and liver cirrhosis as predictive features, clearly aligning with clinical expertise and literature find-ings ^34,38,40,49,54^. Still, certain features contributed less than expected to the model feature importance, indicating present biases and calling for cautious interpretation. E.g., the social desirability bias could explain systematic underestimation of the relevance of alcohol consumption in ML models, as representation of alcohol consumption has been reported to be incorrect ^55^. Further, training on populations with better representation of liver cirrhosis as ICD-code could have assigned higher feature importance to liver cirrhosis.

Our study has several limitations. We did not benchmark our models against previously published ML models for HCC risk, most of which were developed in smaller hospital-based cohorts and lacked external testing ^11–13,56^. This was not possible, as none of the prior studies made their model architectures or weights publicly available, highlighting the need for a shift in thinking towards a more collaborative framework. The development of clear thresholds poses a further limitation to our study. The thresholds developed during five-fold cross-validation on UKB training data achieved very high accuracy when validated on the UKB test set comprised of completely independent test centers from different countries. However, it is still biased by the same active recruitment method, resulting in the well characterized reduced disease burden in the overall UKB population ^22^. The All Of Us Research program, however, encompasses mostly hospital data. Therefore, while the model performance is not inferior in AOU vs UKB, the absolute prediction values are higher in AOU, challenging the proposed thresholds and reflecting well-characterized differences in disease burden ^23^. Most-likely, thresholds will have to be developed independently for certain populations, taking into account the respective disease burden, but also economic considerations that might differ depending on respective populations and healthcare systems. Cost-benefit analysis will have to be performed to estimate ideal proportions and screening intervals. Importantly, the number needed to screen (NNS) correlates inversely with the fraction of potential HCCs one will detect. Aiming at a detection rate of 2 out of 3 of cases of HCC in the screening program (reflecting our rule-in threshold of 0.55) would lead to a NNS of 70-80 in the general population, or a NNS of 26 in the PAR population, with higher or lower NNS according to the desired detection rate (Supplementary Table 19, 21). In contrast to this, guideline-adhering screening programs could have only detected 1 in 5 HCC cases even in the unlikely scenario of perfect screening attendance and perfect detection rate (Figure 2d).

In clinical routine, estimation of annual risk bears greater clinical relevance compared to all-time risk of HCC. Our study lacks models of individual changes in risk of HCC due to lack of trajectory data. In future work, coupling time-to-event analysis with ML-decision trees such as random survival forests, could be beneficial ^57^. Our analysis, like all population-based studies, relies on accurate encoding of EHR. Notably, evaluating our models exclusively on cases with HCC encoding both in NHS and national cancer registries, opposed to NHS-only, revealed a strongly reduced number of false negatives as opposed to evaluation on all EHR-encoded cases. This suggests that those cases, initially labeled as FN, were actually TN and the model was correct when interpreting low-risk phenotypes as negative. Still, a fraction of approximately 5-10 % of HCC cases were wrongly assigned to the low-risk group, for UKB and AOU test sets alike. Analysis of these misclassified cases revealed notable insights. Consistently, our classifiers demonstrated a higher tendency to overlook HCC in female patients compared to male patients. This could be explained by an overall less prominent HCC phenotype in females or the higher prevalence for males, challenging the assumption that a combined model for all sexes is actually feasible. Interestingly, this performance gap was less-pronounced in the All Of Us cohort, with furthermore no differences in performance for the “White” ethnicity vs other ethnicities, suggesting good generalizability. However, validation in other populations, especially Asian populations with higher prevalence of viral hepatitis, is still pending.

There are numerous obstacles on the path to clinical implementation of ML risk models for HCC prediction. Firstly, prospective clinical trials will be needed before clinical adoption can be recommended. Setting those up isolated for HCC-screening will be challenging due to its low incidence. We show that in most cases, steatotic liver diseases are only diagnosed after diagnosis of HCC. This suggests that current standard practice, which links HCC screening to prior diagnosis of liver cirrhosis, is likely too narrowly defined, and that accurate, non-invasive biomarkers are needed both for prediction of HCC and for prediction of cirrhotic liver disease. It could therefore be beneficial to set up multi-purpose trials that screen for liver morbidity in general and HCC alike, as e.g. the LiverAIM trial aims to do ^58^. Current clinical routine demands an incidence of 1.5 % in the screening group for HCC screening to become economically feasible ^59^, and a minimal annual incidence of 0.2 % in the respective population to allow for effective screening ^7,48^. Patients in our model’s high-risk group clearly surpass the annual incidence of 0.2 % in both cohorts, even despite removal of HCC cases near baseline, removal of cases not confirmed by cancer registries, and a lower general incidence of HCC in the UKB than in the general population. While thresholds would have to be adapted to surpass the limit of 1.5 % incidence, this limit might change in favor of broader screening strategies, as treatment with expensive immune checkpoint inhibition proves successful also in earlier stages, increasing therapy cost extensively ^60^.

From a technical perspective, our models can be integrated effortlessly for further research purposes and, once a medical device, in clinical practice. The simple random forest architecture with as few as 15 affordable and routinely available parameters can be validated on independent data with the python package we provide. Further, for research-purposes only, it can be accessed for individual inference via a web-tool we provide at https://hugging-face.co/spaces/schneiderlab/ML-HCC, which also agentic workflows using large-language-models could access in the future ^61^. This could enable inference of a multitude of prediction models at once compared to tedious manual variable entering, as the latter will naturally limit clinical usage ^61^. Finally, long-term integration should ideally happen interoperable with hospi-tal-information-systems, where input data can be automatically inferred for a multitude of models, as machine-learning predictions enter clinical routine in virtually every discipline of healthcare ^62,63^.

Our study reveals critical gaps in the current screening process for HCC. It addresses a clinically highly relevant question in an interpretable way with actionable output. Our model out-performs all established linear risk calculators for HCC by leveraging interpretable machine learning and extensive population-based data. As risk calculators are a routinely used practice especially in hepatology, e.g. for estimation of liver-related mortality ^64^, its implementation into clinical routine could come naturally, especially given its reliance on simple, ubiquitously available parameters. We are confident that, after necessary prospective evaluation, our model can therefore contribute as a decision-aid for clinicians, integrating risk factors for a personalized assessment of risk, allowing earlier intervention and potentially curative treatment.

## Supporting information

Supplementary Material (except Tables)

## Data Availability

UK Biobank data, including NMR metabolomics, are publicly available to bona fide researchers upon application at http://www.ukbiobank.ac.uk/using-the-resource/. Detailed information on predictors and endpoints used in this study is presented in Supplementary Tables 1-25. This study used data from the All of Us Research Program's Controlled Tier Dataset v7, available to authorized users on the Researcher Workbench.

https://allofus.nih.gov/

## Abbreviations

AASLD: American Association for the Study of Liver Diseases
ALT: Alanine Aminotransferase
AOU: All Of Us Research Program
AST: Aspartate Aminotransferase
AUPRC: Area Under the Precision-Recall Curve
AUROC: Area Under the Receiver Operating Characteristic Curve
BMI: Body Mass Index
CI: Confidence Interval
CLD: Chronic Liver Disease
COPE: Committee on Publication Ethics
EASL: European Association for the Study of the Liver
EHR: Electronic Health Records
FDR: False Discovery Rate
FN: False Negative
FP: False Positive
γ-GT: Gamma Glutamyltransferase
HCC: Hepatocellular Carcinoma
ICD: International Classification of Diseases
IGF-1: Insulin-like Growth Factor 1
MASLD: Metabolic Dysfunction-Associated Steatotic Liver Disease
ML: Machine Learning
NMR: Nuclear Magnetic Resonance
NNS: Number Needed to Screen
NPV: Negative Predictive Value
OMOP: Observational Medical Outcomes Partnership
PAR: Patients at Risk
PPV: Positive Predictive Value
PRC: Precision-Recall Curve
PRS: Polygenic Risk Score
RFC: Random Forest Classifier
ROC: Receiver Operating Characteristic
SD: Standard Deviation
SHAP: SHapley Additive exPlanations
SNP: Single Nucleotide Polymorphism
TN: True Negative
TP: True Positive
TRIPOD: Transparent Reporting of a multivariable prediction model for Individual Prognosis Or Diagnosis
UKB: UK Biobank
XGB: Extreme Gradient Boosting

## Additional Information

## Acknowledgements

This research has been conducted using the UK Biobank Resource under Application Number 71300. UK biobank data was accessed by J.N.C, C.V.S and K.M.S. Copyright © 2024, NHS England. Re-used with the permission of the NHS England and/or UK Biobank. All rights reserved. This work uses data provided by patients and collected by the NHS as part of their care and support. All UKB analyses have been performed by JC and PHK. The All of Us Research Program is supported by the National Institutes of Health, Office of the Director: Regional Medical Centers (OT2 OD026549; OT2 OD026554; OT2 OD026557; OT2 OD026556; OT2 OD026550; OT2 OD 026552; OT2 OD026553; OT2 OD026548; OT2 OD026551; OT2 OD026555); Inter agency agreement AOD 16037; Federally Qualified Health Centers HHSN 263201600085U; Data and Research Center: U2C OD023196; Genome Centers (OT2 OD002748; OT2 OD002750; OT2 OD002751); Biobank: U24 OD023121; The Participant Center: U24 OD023176; Participant Technology Systems Center: U24 OD023163; Communications and Engagement: OT2 OD023205; OT2 OD023206; and Community Partners (OT2 OD025277; OT2 OD025315; OT2 OD025337; OT2 OD025276). We gratefully acknowledge All of Us participants for their contributions, without whom this research would not have been possible. We also thank the National Institutes of Health’s All of Us Research Program for making available the participant data examined in this study.

## Author contributions

JC, JNK and CVS conceptualized the study and wrote the manuscript. JC and FvH executed the preprocessing. JC and PHK built the modeling pipeline. JC and DZ executed the external validation. PHK and TS built the web-application. OSMEN, LZ, JuC, FT, TL, TB, AK, DT and KMS provided invaluable feedback and revised the manu-script.

## Code availability

All code developed and used throughout this study has been made open source and is available on GitHub. The code used for preprocessing can be found here: Code for model development can be found here: https://github.com/schneiderlabac/hcc_u_soon

## Declaration of generative AI and AI-assisted technologies in the writing process

During the preparation of this work the authors used GPT-4o (OpenAI) and Claude 3.5 Sonnet (Anthropic) in order to correct spelling and grammar and for coding assistance, in accordance with the COPE (Committee on Publication Ethics) position statement of 13 February 2023 ^67^. After using this tool/service, the authors reviewed and edited

## Ethics disclaimer

We acknowledge the ethical complexities of categorizing individuals in medical research, especially when categorizing by ethnicity. While any form of categorization risks perpetuating discrimination, the complete omission of such considerations in medical AI could paradoxically reinforce healthcare disparities. Machine learning models are sensitive to training data composition and can silently perpetuate or amplify existing biases if their performance across different populations remains untested. By explicitly examining model performance across ethnic groups, we aim to ensure equitable predictive accuracy and identify potential disparities in model generalizability that require attention. This approach aligns with the broader goal of developing inclusive healthcare solutions while remaining mindful of the need to handle demographic data with appropriate sensitivity and scientific rigor. Our categorizations follow standardized reporting guidelines while recognizing that such classifications are social constructs and cannot fully capture human diversity. Our study does not include confidential information. All research procedures were conducted exclusively on anonymized patient data and in accordance with the Declaration of Helsinki, maintaining all relevant ethical standards.

