## Supplementary Material (except Tables) for "Machine learning predicts liver cancer risk from routine clinical data: a large population-based multicentric study"

#### Table of Contents:

**Supplementary Figures** **2**
- Supplementary Fig. 1: Data processing flowchart 2
- Supplementary Fig. 2: Preprocessing of study participants. 3
- Supplementary Fig. 3: Additional performance metrics 4

Table of content for supplementary tables 5

### Supplementary Figures

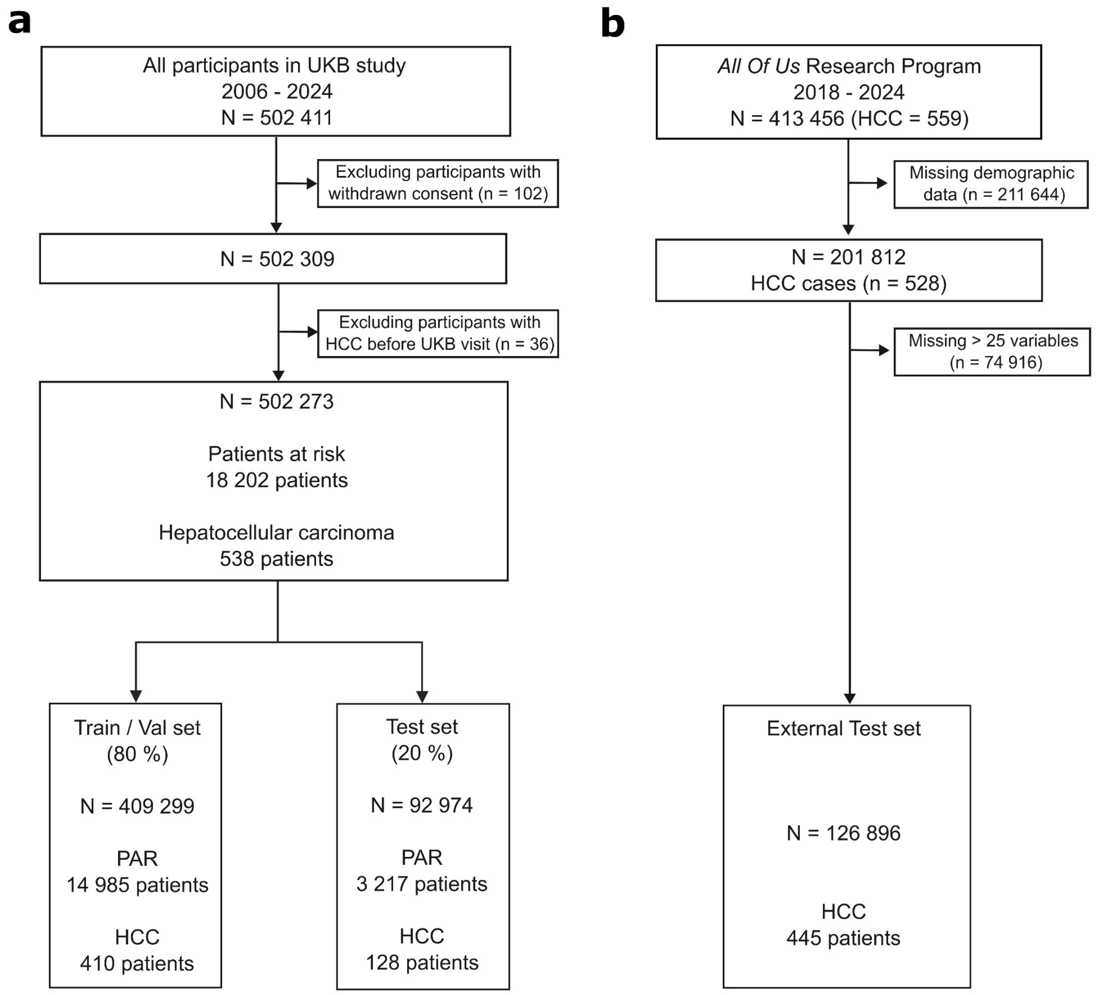


**Supplementary Fig. 1: Data processing flowchart**

**a** Data processing flowchart for UKB participants. Val = Validation. **b** Data processing flowchart for participants in "All Of Us" Research Program

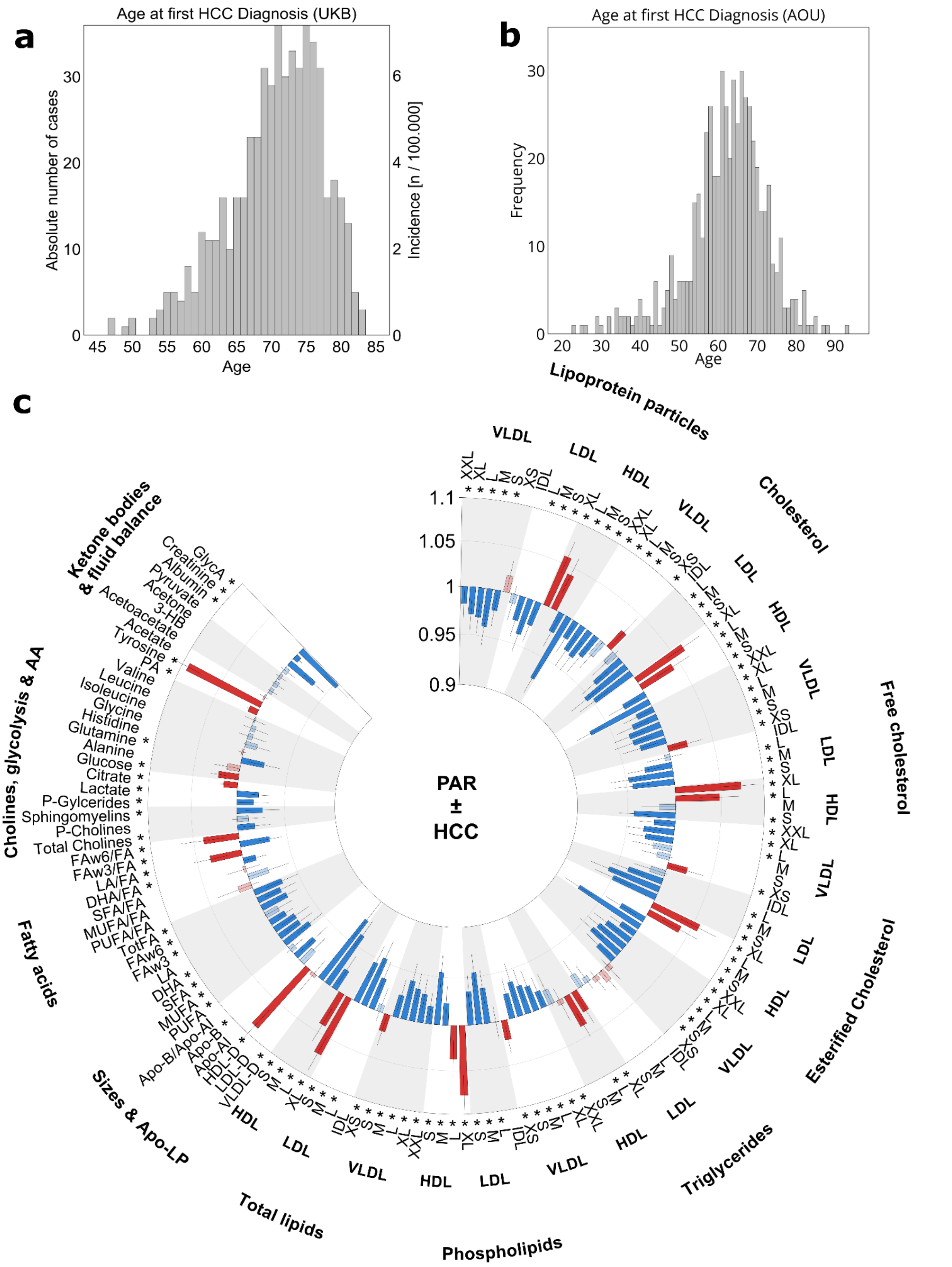


**Supplementary Fig. 2: Preprocessing of study participants.**

**a, b** Age in years of participants at first-documented diagnosis of HCC. **a** for UKB and **b** for All Of Us Research Program. **c** Associations of metabolic biomarkers with development of HCC. Hazard ratios + 95% confidence intervals as 1-SD higher metabolic biomarker on the natural log scale, stratified by age, sex, BMI, compared to cohort with pre-diagnosed liver disease or elevated liver enzymes, jointly defined as patients-at-risk (PAR). *False discovery rate-controlled $p < 0.01$.

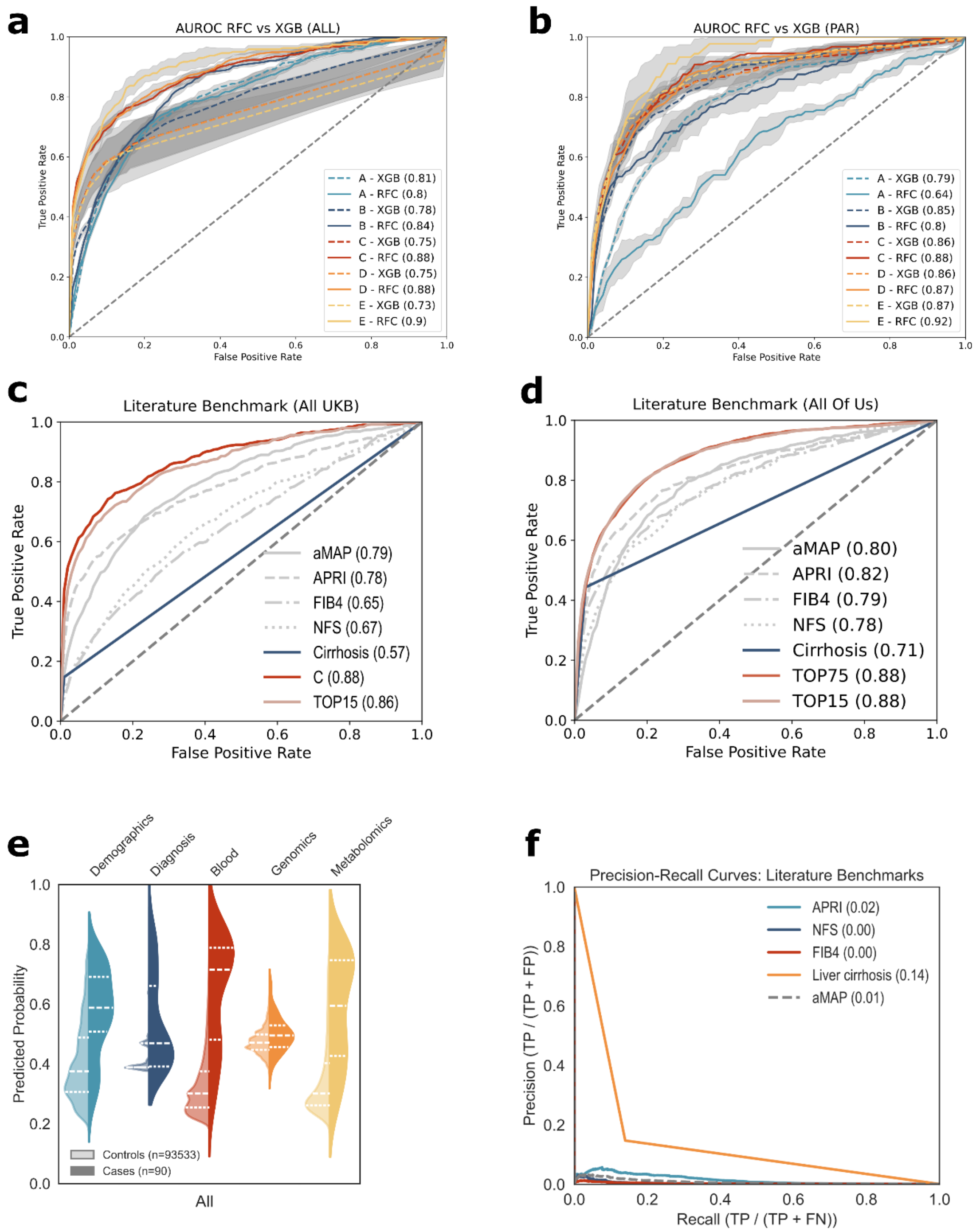


**Supplementary Fig. 3: Additional performance metrics**

**a, b** Model performance on the test set of UKB as receiver operating characteristic (ROC) curves and corresponding area under the curve (AUC) for either all participants (a) or patients with pre-diagnosed liver disease (b). Each line represents the performance of one majority vote model from five-fold cross-validation ± standard deviation of true positive rate, either for the random forest classifier (straight line) or extreme gradient boost (dotted line). **c** Performance of various established linear risk scores, cirrhosis as binary marker for all of UKB (England, Scotland, Wales), plotted against model performance of newly developed Model C and Model TOP15 for UKB test set performance (Scotland, Wales). **d** Risk scores as in c, but for AOU cohort. Models TOP75 and TOP15 applied to the whole AOU cohort. **e** Split violin distributions for prediction scores of separate models per modality for UKB cohort "All". **f** Precision-recall curves for literature benchmarks in UKB "All" cohort TP = True Positives, FP = False Positives, FN= False Negatives.

#### Table of content for supplementary tables (see Supplementary_Material.xlsx)

| Table | Description | Cohort |
|---|---|---|
| Supplementary Table 1 | ICD10 Codes used for Patients at risk definition | UKB/AOU |
| Supplementary Table 2 | ICD10 Codes defined as single-entity feature | UKB/AOU |
| Supplementary Table 3 | ICD10 Codes defined as grouped feature | UKB/AOU |
| Supplementary Table 4 | Assessment Features | UKB |
| Supplementary Table 5 | Blood Count Features | UKB/AOU |
| Supplementary Table 6 | Metabolomics Features | UKB |
| Supplementary Table 7 | Single Nucleotide Polymorphisms Data | UKB |
| Supplementary Table 8 | Mapping for self-reported diagnosis codes | UKB/AOU |
| Supplementary Table 9 | At-risk subsets and respective HCC/control cohort sizes | UKB |
| Supplementary Table 10 | Center splits | UKB |
| Supplementary Table 11 | HCC cases per center | UKB |
| Supplementary Table 12 | Mappings UKB_AOU MinMax Median Mean and Conversion | UKB/AOU |
| Supplementary Table 13 | Table ICD | UKB |
| Supplementary Table 14 | Table ICD | AOU |
| Supplementary Table 15 | Table Blood Count/Biochemistry stratified for HCC and SEX | UKB |
| Supplementary Table 16 | Table Blood Count/Biochemistry stratified for HCC and SEX | AOU |
| Supplementary Table 17 | Metabolomics Stratified for HCC | UKB |
| Supplementary Table 18 | Model Performances threshold-independent (AUROCs, AUPRCs) | UKB |

| | | |
|---|---|---|
| Supplementary Table 19 | Model performances threshold-dependent | UKB |
| Supplementary Table 20 | Model Performances threshold-independent (AUROCs, AUPRCs) | AOU |
| Supplementary Table 21 | Model performances threshold-dependent | AOU |
| Supplementary Table 22 | Prediction Value Comparison | UKB/AOU |
| Supplementary Table 23 | Delong Tests | UKB |
| Supplementary Table 24 | Delong Tests | AOU |
| Supplementary Table 25 | Feature Importances Model A-E + TOP75-TOP15 UKB | UKB |
